# Antibiotic-resistance plasmid amplified among MRSA cases in an urban jail and its connected communities

**DOI:** 10.1101/2025.10.23.25338646

**Authors:** Stephanie N. Thiede, Hannah Steinberg, Emily Benedict, Sarah Sansom, Alla Aroutcheva, Kyle J. Gontjes, Katherine Winner, Sonya Royzenblat, Ali Pirani, Robert A Weinstein, Chad Zawitz, Nicholas M. Moore, Kyle J. Popovich, Evan S. Snitkin

## Abstract

Jails have been hypothesized to be hotspots for the spread of methicillin-resistant *Staphylococcus aureus* (MRSA). We integrate genomic and epidemiologic data to investigate USA300 MRSA transmission in an urban jail and its connected communities. A genome-wide association study of 308 jail isolates from 2015-2018 revealed a plasmid encoding the *ermC* clindamycin/erythromycin resistance gene was associated with a 6-fold increased odds of MRSA genetic linkages among detainees. Additionally, 52% of jail-onset MRSA infections carried this plasmid compared to 14% of intake colonization isolates, supporting its role in MRSA spread in the jail. Extending our analysis to 774 isolates from a local healthcare system from 2011-2014, the *ermC-*carrying plasmid was also associated with MRSA transmission in the larger community and was enriched among former jail detainees and those with related isolates to recently incarcerated cases. Lastly, topical clindamycin exposure before MRSA infection was associated with *ermC* plasmid presence in both settings, but exposure prevalence was higher in jail versus community cases (7.5% vs. 0.9%), suggesting antibiotic use in the jail may have created a favorable environment for the spread of *ermC*-carrying strains. These findings highlight the impact of antibiotic use in jails on antibiotic resistance in both jails and their surrounding communities.

## Introduction

Once primarily a healthcare-associated pathogen, starting in the late 1990’s and early 2000’s methicillin-resistant *Staphylococcus aureus* (MRSA) transmission began occurring in community settings(1). In the United States, this transition was largely due to the emergence of the USA300 strain of MRSA, a sub-lineage of the ST8/CC8 clonal complex(2). Initially, USA300 outbreaks were observed in congregate settings defined by close personal contact, such as jails, prisons, military barracks, daycares, and gyms(3). Subsequently, USA300 has become endemic in many communities, and while rates of healthcare-associated MRSA infections have declined with enhanced prevention efforts, community-associated MRSA (CA-MRSA) infection rates have largely remained stable(4).

USA300 has been characterized by higher rates of susceptibility to non-beta-lactam antibiotics in comparison to more typical healthcare-associated MRSA strains(5, 6). This feature has directly impacted outpatient management of skin and skin structure infections due to CA-MRSA by allowing oral antibiotic options as empiric therapy. However, resistance in CA-MRSA strains has steadily emerged, with notable variation in resistance to agents such as fluoroquinolones and clindamycin across different regions of the US(7–9). Furthermore, multi-drug resistant USA300 MRSA has been observed in several US cities(7, 8), highlighting the importance of antibiotic stewardship efforts both in and out of healthcare settings and for continued monitoring of local antibiograms for CA-MRSA strains. Importantly, due to lack of regular genomic surveillance, shifts in circulating lineages and resistance determinants that are driven by local selective pressures likely go undetected (10).

As the community MRSA epidemic has progressed and become endemic, significant disparities have arisen with respect to those communities most impacted. Previous studies in Chicago, IL have found that the zip codes with the highest rates of CA-MRSA are those with low socio-economic status, high rates of unstable housing, high rates of substance abuse, and high rates of detainee release from correctional facilities(11–15). Jails have garnered particular interest as potential amplifiers of community MRSA because of high rates of community influx, recidivism, and the potential for intermixing of individuals from different social networks and communities(16). Previous mathematical modeling studies have provided support for the potential importance of jails as amplifiers of MRSA, with the number of contacts during incarceration and inflow of infected individuals playing a significant role in spread(17). We previously found a high colonization prevalence for MRSA among both female (20%)(18) and male (19%)(19) detainees when entering the jail, with genomic analysis supporting the spread of MRSA during incarceration(20). We also found that incarceration rates are a key driver of community MRSA rates and racial disparities in infections at the census-tract level (15). However, the mechanism by which MRSA dynamics within urban jails impact the surrounding community remain unclear. Determining the relative influence of jail and community transmission on CA-MRSA incidence is essential to determining where targeted surveillance and intervention efforts will be most impactful.

Here, we sought to improve our understanding of the intersection between community and jail transmission networks by leveraging large genomic and epidemiologic data sets for individuals colonized or infected with USA300 MRSA at Cook County Jail (CCJ), and Cook County Health (CCH), a safety-net healthcare network serving communities with high detainee release rates. By employing genome-wide association studies (GWAS) to identify genotypes preferentially spreading in jail, and integrating clinical metadata, we find evidence for the amplification of an *ermC*-carrying resistance plasmid via clonal spread of multiple USA300 sub-lineages that independently acquired the *ermC* plasmid, potentially favored by high rates of topical clindamycin use among detainees during the study period. GWAS of CCH isolates revealed proliferation of the same plasmid, with direct or indirect exposure to CCJ being associated with having a MRSA strain that harbored the *ermC* resistance plasmid. These findings highlight the distributed impact antibiotic use has across community settings, and how the proliferation of CA-MRSA strains within urban jails can leave detectable signatures on communities with high detainee release.

## Results

### Characteristics of MRSA cases at Cook County Jail

From 2015-2018, 308 unique USA300 MRSA isolates were cultured from 305 detainees at CCJ. Three (1%) of these cases were classified as community-onset infections (infection culture obtained less than 72 hours after entering CCJ), 146 (47%) were jail-onset infections, 147 (48%) were intake colonizations, and 12 (3.9%) were jail-onset colonizations. The majority (74%) of cases were male. Colonization isolates were cultured from the nose (n=89), throat (n=40), or groin (n=30) of asymptomatic cases, and the vast majority of symptomatic infections were wound infections (n=145) **(Supplemental Table S1).**

### ermC-carrying plasmid associated with transmission in Cook County Jail

We previously conducted a genomic epidemiology analysis of USA300 MRSA isolates in CCJ and found evidence of within-jail transmission, with a majority of individuals with jail-acquired infections harboring genetic linkage to another detainee’s MRSA isolate, and significant spatiotemporal overlap among linked individuals(20). A striking feature of these putative within-jail transmission linkages is their formation of large clusters ranging in size from 2 to 13 members. These clusters spanned the USA300 phylogeny (**Figure 1**), likely due to the high rate of MRSA importation into the jail (19% colonization prevalence) leading to sampling of USA300 diversity in the community(19). While there are potential epidemiologic explanations for this observation that are consistent with our prior analysis (e.g. environmental reservoirs, super spreaders), we here considered the possibility that there were also microbial genetic factors contributing to the preferential spread of these genomic clusters. To test this hypothesis, we determined the accessory genes present in each USA300 MRSA genome and tested the association with genomic transmission linkage, defined as being related to another isolate within 20 SNVs (20, 21).

**Figure 1:**
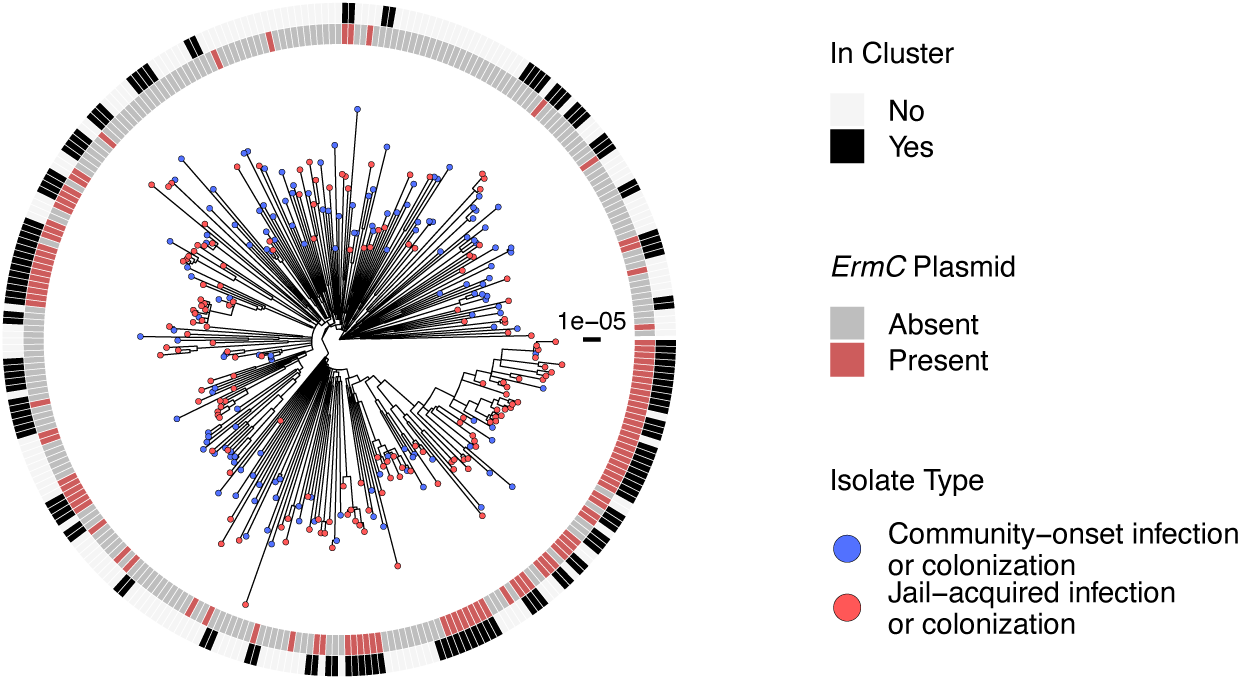
Phylogenetic tree of USA300 MRSA in Cook County Jail. Maximum likelihood phylogeny of jail USA300 samples created with IQTREE using core-genome variants and ultrafast bootstrap with 1000 replicates. Scale bar indicates substitutions per site. Tips designate if the isolate was acquired within the jail (MRSA colonization not observed at intake but observed at 30 days within jail or infection after 72 hours of entering jail, shown in red) or community (MRSA colonization positive at intake or infection within 72 hours of entering jail, shown in blue). Inner ring indicates the presence (red) or absence (grey) of the *ermC-*carrying plasmid. Outer ring indicates if the isolate is genetically related to any other isolate within 20 SNVs (black) or not (white).

There were two genes found to be significantly associated with transmission after multiple test correction (**Figure 2A**). By several orders of magnitude, the most significant associations were *ermC* (OR = 5.56, p = 1.3×10^−11^, 95% CI: 3.30-9.36) and a gene annotated as a replication and maintenance protein (OR = 5.96, p = 2.15×10^−12^, 95% CI: 3.52-10.09), with these genes having > 90% co-occurrence across genomes. Comparison to sequence databases revealed that these genes are part of a small, two-gene 2.4 kb plasmid that confers resistance to macrolides and lincosamides and has been previously observed in USA300 MRSA(22, 23). We confirmed the presence of this plasmid by mapping reads from MRSA genomes to the plasmid sequence (see **Methods**). Comparing the pangenome presence/absence of *ermC* and replication and maintenance gene to the plasmid presence resulted in 98% concordance (See **Supplemental Table S2**). Examining antibiotic susceptibility data for CCJ MRSA isolates supports most plasmids conferring resistance to both erythromycin and clindamycin, with 55 of the 60 plasmid harboring isolates with available antibiotic susceptibility data being resistant to both antibiotics, and the remaining 5 being resistant to only erythromycin(24). Lastly, we noted that the *ermC*-carrying plasmid was present in distinct clades across the USA300 phylogeny, indicating that this plasmid was acquired multiple times and subsequently spread (**Figure 1**).

**Figure 2:**
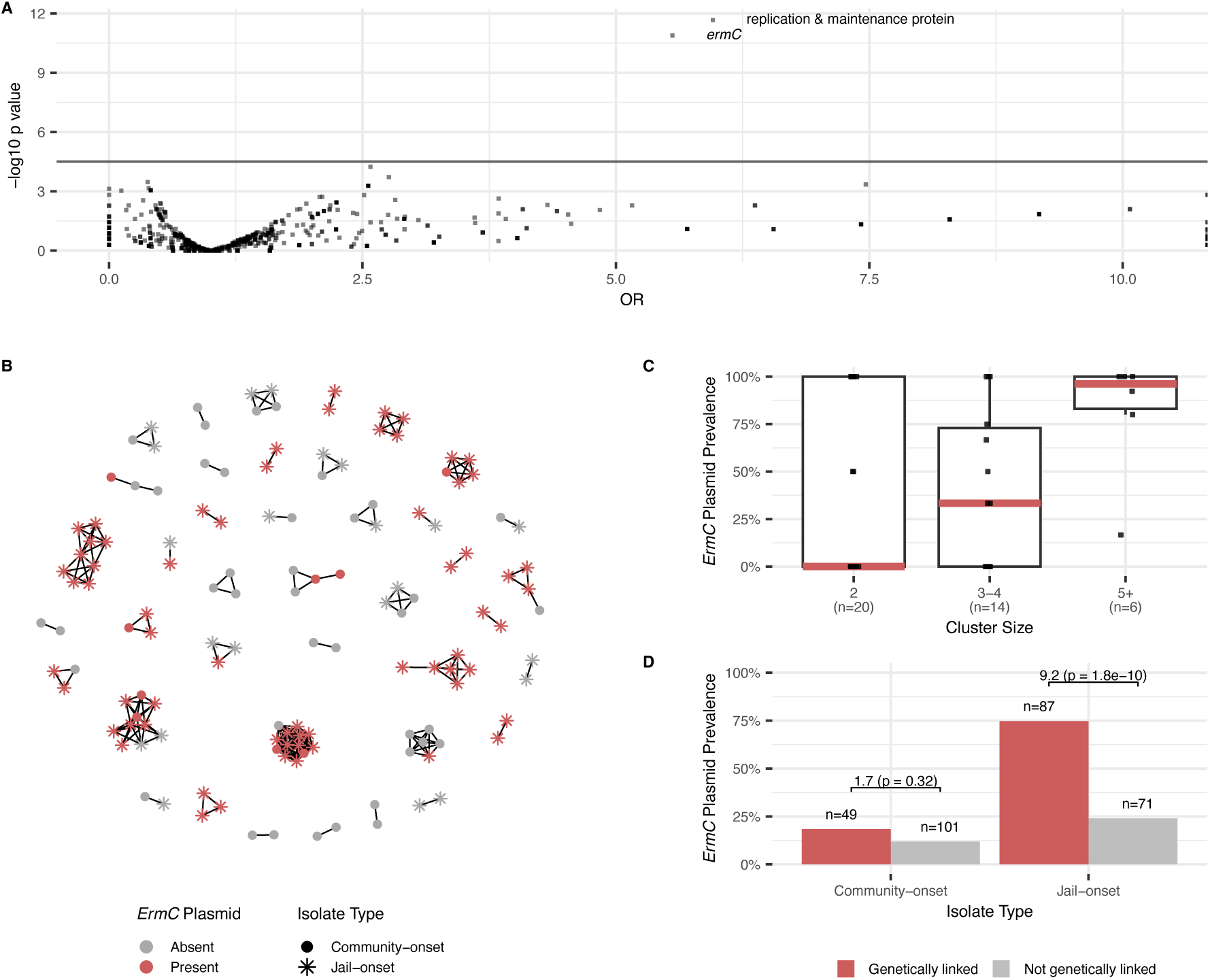
Association between pangenome and transmission in the Cook County Jail from 2015-2018. **A)** A genome-wide association study was performed to measure the association between accessory gene presence and genetic linkage to another CCJ isolate within 20 SNVs. P-values represent unadjusted p-values from two-sided Fisher’s exact tests. Horizontal line indicates Bonferonni adjusted p-value threshold. **B)** Network diagram of MRSA transmission clusters. Edges indicate that isolates are related within 20 SNVs. Nodes colored by presence (red) or absence (grey) of the *ermC*-carrying plasmid in the corresponding isolate. Shape indicates the isolate type, where stars are jail-acquired colonization or infections (MRSA colonization not observed at intake but observed at 30 days within jail or infection after 72 hours of entering jail) and circles are community-acquired colonization or infection (MRSA colonization positive at intake or infection within 72 hours of entering jail). There were 172 unclustered isolates in our analysis. Of these, 29 (16.9%) had the *ermC* plasmid present, 101 (58.7%) were community-onset cases, and 71 (41.3%) were jail-onset cases. Of the unclustered community-onset cases, 12 (11.9%) carried the *ermC* plasmid, while 17 (23.9%) of the unclustered jail-onset cases had the *ermC* plasmid present. **C)** Larger clusters of MRSA isolates are associated with higher *ermC* prevalence than small clusters. Boxes represent the interquartile range (IQR) of *ermC* plasmid prevalence, median *ermC* plasmid prevalence is represented by red horizontal lines, and whiskers represent the range of *ermC* plasmid prevalence in each cluster size group (clusters of 2 isolates, 3-4 isolates, or 5 or more isolates) excluding outliers (values more than 1.5 times the IQR from the 25th or 75th percentile for a given group). The number of clusters in each size category are indicated under the x-axis labels. **D)** Bars represent the proportion of isolates carrying the *ermC* plasmid in community-onset and jail-onset MRSA colonization and infections by whether the case was genetically linked to another case in the jail (red) or not (grey). The presence of the *erm*C plasmid is associated with genetic linkage at CCJ, but not statistically significant when considering only surveillance for colonization on entrance to CCJ (first bars). Numbers above bars represent sample sizes, and odds ratios and p-values comparing *ermC* prevalence by genetic linkage were determined using two-sided Fisher’s exact tests.

Examining the presence of *ermC* on the USA300 phylogeny shows its concordance with the large clusters of MRSA transmission previously observed in the jail (**Figure 1**). Twenty-two out of the 40 identified MRSA transmission clusters in the jail contained at least one isolate with the *ermC* plasmid (**Supplemental Table S2**). Further, we observed that individuals harboring MRSA with the *ermC*-carrying plasmid were part of significantly larger transmission clusters, with jail clusters comprised of 5 or more isolates having 2.4 times (Poisson univariate regression; 95% CI: 1.3-4.4; p = 0.005) increased prevalence of *erm*C as clusters with 2 people (**Figures 2B and 2C**), further implicating the role of the plasmid in MRSA proliferation within the jail. Additional support for the amplification of *ermC* within the jail comes from comparing the prevalence of *ermC* among admission versus acquired isolates. In particular, the prevalence of isolates carrying this plasmid was significantly higher among jail-acquired infections than among isolates present at intake to the jail (52% vs. 14% respectively, p =8.95 × 10^−13^). Taken together, these results indicate that there were multiple importations of *ermC* harboring strains into the jail, and that these preferentially spread among detainees as compared to strains not harboring *ermC*. However, we also noted that despite the lower prevalence of *ermC* on admission to CCJ, intake positive isolates with the *ermC*-carrying plasmid were still more often genetically linked to other intake isolates than non-*ermC* strains, though not significantly so (OR = 1.7, p = 0.32, 95% CI: 0.58-4.3), potentially reflective of preferential spread of plasmid carrying strains in the broader community (**Figure 2D** and **Supplemental Figure S1**).

### ermC-carrying plasmid is associated with transmission in the larger community served by Cook County Health

To further explore the potential role of the *ermC*-carrying plasmid in transmission outside of the jail, we leveraged a previously sequenced set of 774 genomes comprising a comprehensive collection of clinical cultures from patients at Cook County Health (CCH) collected from 2011-2014(25). The communities served by CCH overlap with several zip codes identified to be areas of high detainee release from correctional facilities (12, 13, 15). Moreover, our prior genomic epidemiology analysis of the CCH isolates supports community origins for even healthcare-onset infections(25). Thus, we hypothesized that this collection from CCH would provide insight into MRSA circulation in the communities most connected to the jail.

To evaluate whether *ermC* was also associated with transmission in the CCH collection, we performed an independent GWAS to identify accessory genes associated with genomic transmission linkage within the CCH collection. While several additional accessory genes were found to be associated with transmission, by far the most significantly associated genes were the two *ermC* plasmid genes (OR = 3.62 and 3.74, p = 1.24×10^−10^ and 3.14×10^−12^, 95% CI: 2.46-5.36 and 2.57-5.43 for *ermC* and replication and maintenance protein, respectively, **Supplemental Figure S2**). As with MRSA from CCJ, antibiotic susceptibility data supported most CCH MRSA strains harboring the plasmid that conferred resistance to both clindamycin and erythromycin, with 132 of 162 strains with the *ermC* plasmid being resistant to both antibiotics, and the other 30 being resistant to only erythromycin. As with the CCJ isolates, concordance between pangenome presence/absence and plasmid presence was high (94% concordance). From here forward, the presence of the plasmid based on read mapping to USA300-SUR4 was used as a surrogate for tracking *ermC* presence (see **Methods**).

Also like the CCJ collection, the presence of *ermC* was a result of multiple acquisitions of the plasmid among isolates spanning the USA300 phylogeny (**Supplemental Figures S3 and S4**). Using an ancestral reconstruction approach we estimate a total of 91 independent plasmid acquisition events across the combined CCH/CCJ USA300 phylogeny (**Supplemental Figure S5**), which vary in their magnitude of subsequent transmission (1 – 60 individuals) and their persistence over time (2 – 2,413 days). As with CCJ, harboring *ermC* in the CCH collection was associated with larger transmission clusters than among isolates not harboring the plasmid-*ermC* plasmid prevalence was 1.9 times greater in clusters with five or more individuals than in clusters with 2-4 individuals (Poisson univariate regression; 95% CI: 1.2-3.1; p = 0.005). (**Supplemental Figure S2**). Further supporting an advantage for *ermC* in the broader community, we observed that strains harboring the *ermC* plasmid increased in prevalence over time from 2011-2014 (Χ^2^ test for trend = 4.1, p = 0.04) (**Supplemental Figure S2**). Moreover, taking advantage of repeat clinical cultures for 11 patients, we observed additional evidence that amplification of *ermC* in the community is not just driven by transmission, but also continued horizontal transfer into new genetic backgrounds (**Supplemental Figure S6**). In particular, we observed two cases (Patients D and H) where patient’s repeat culture isolates were within 20 single nucleotide variants (SNVs), and the *ermC* plasmid was not present in the first clinical culture, but was present in the second, consistent with acquisition of the plasmid by horizontal gene transfer.

### Clindamycin use in the Cook County Health and Jail populations associated with ermC

The observation of an *ermC* plasmid conferring resistance to clindamycin being associated with sustained transmission over time was at odds with prior work suggesting a significant fitness cost (26). We hypothesized that one pathway for preferential spread of *ermC* plasmids was high rates of clindamycin use. Indeed, among patients represented in the CCH collection, clindamycin use was significantly associated with having a MRSA isolate harboring *ermC* (**Figure 3A**, **Table 1**, OR = 3.1, p = 6.5×10^−6^, 95% CI: 1.9-5.0). Moreover, the association between harboring an *ermC* strain was even stronger with topical clindamycin exposure (**Figure 3A**, OR = 27, p = 2.3×10^−4^, 95% CI: 3.7-621). Similarly, among detainees with jail-onset MRSA infections at CCJ, *ermC* was associated with both overall clindamycin exposure (OR = 4.5, p = 0.0011, 95% CI: 1.7-12.0) and topical clindamycin exposure (OR = 9.2, p= 0.013, 95% CI: 1.2-202)(**Figure 3A, Supplemental Table S4**).

**Figure 3:**
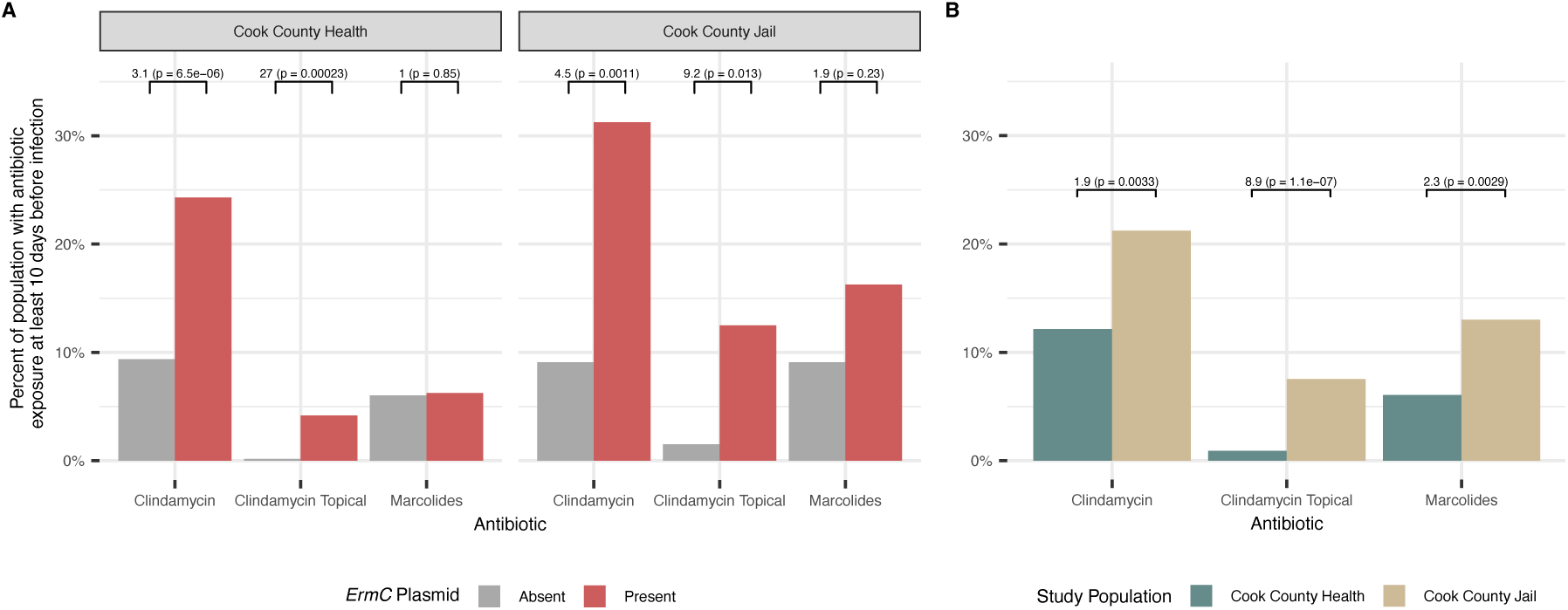
Antibiotic exposure and presence of *ermC*. Antibiotic exposure was defined as at least 10 days prior to infection, to ensure that the antibiotic was not prescribed for the infection, and no more than 6 months prior to infection. (**A)** Bars represent antibiotic exposure by antibiotic type and *ermC* plasmid presence (present: red, absent: grey). Exposure to clindamycin (especially topical) is associated with presence of ermC-carrying plasmid at CCJ and CCH. Odds ratios comparing antibiotic exposure and *ermC* plasmid presence and corresponding p-values (two-sided Fisher’s exact test) are indicated above each location/antibiotic group. **B)** Bars represent antibiotic exposure in MRSA cases by location (CCH: teal, CCJ: tan). Total clindamycin, topical clindamycin, and macrolide use is significantly higher among individuals with MRSA at CCJ, as compared to CCH. Odds ratios comparing antibiotic exposure between CCJ and CCH MRSA cases and corresponding p-values (two-sided Fisher’s exact test) are indicated above each antibiotic group.

**Table 1:**
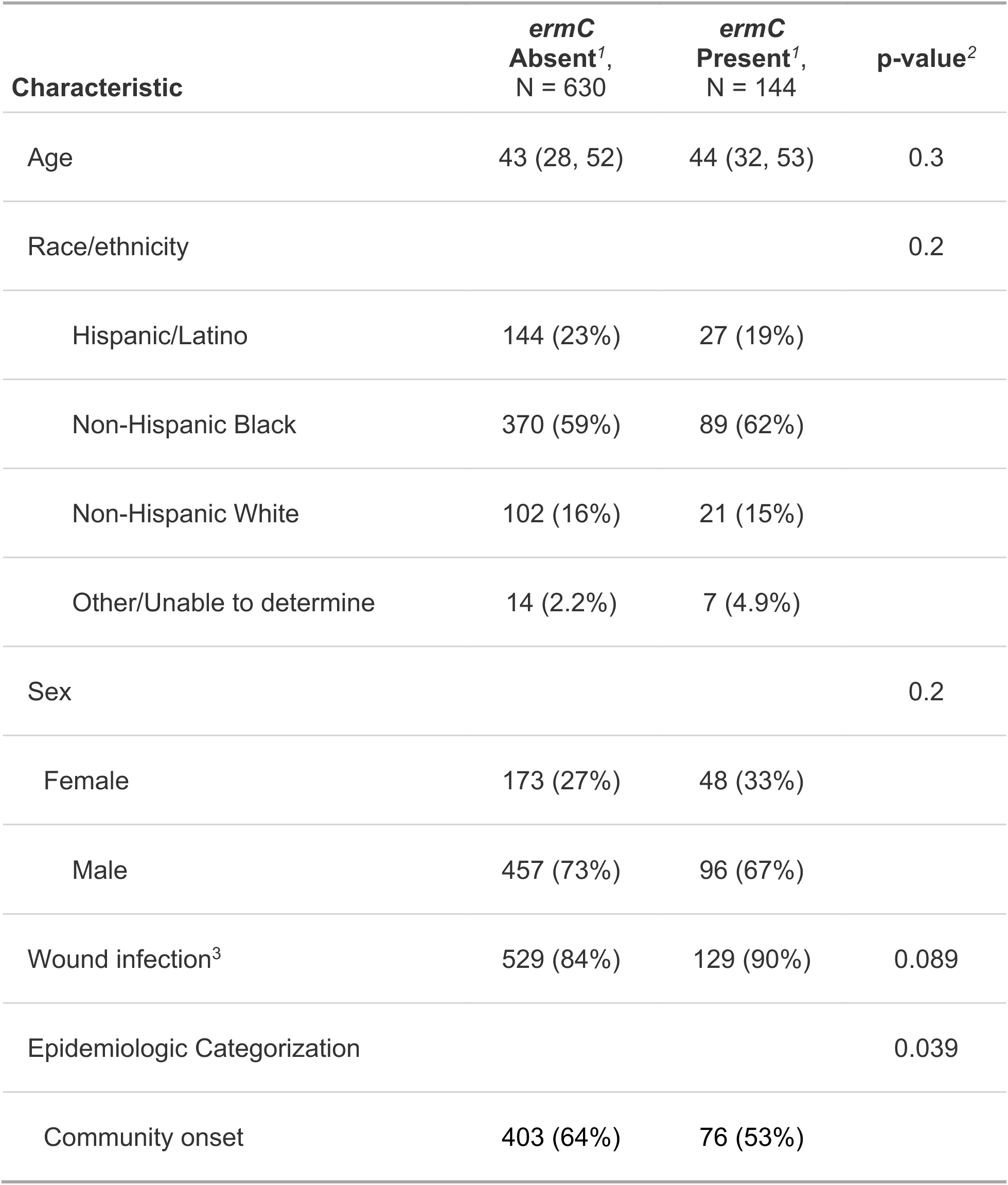
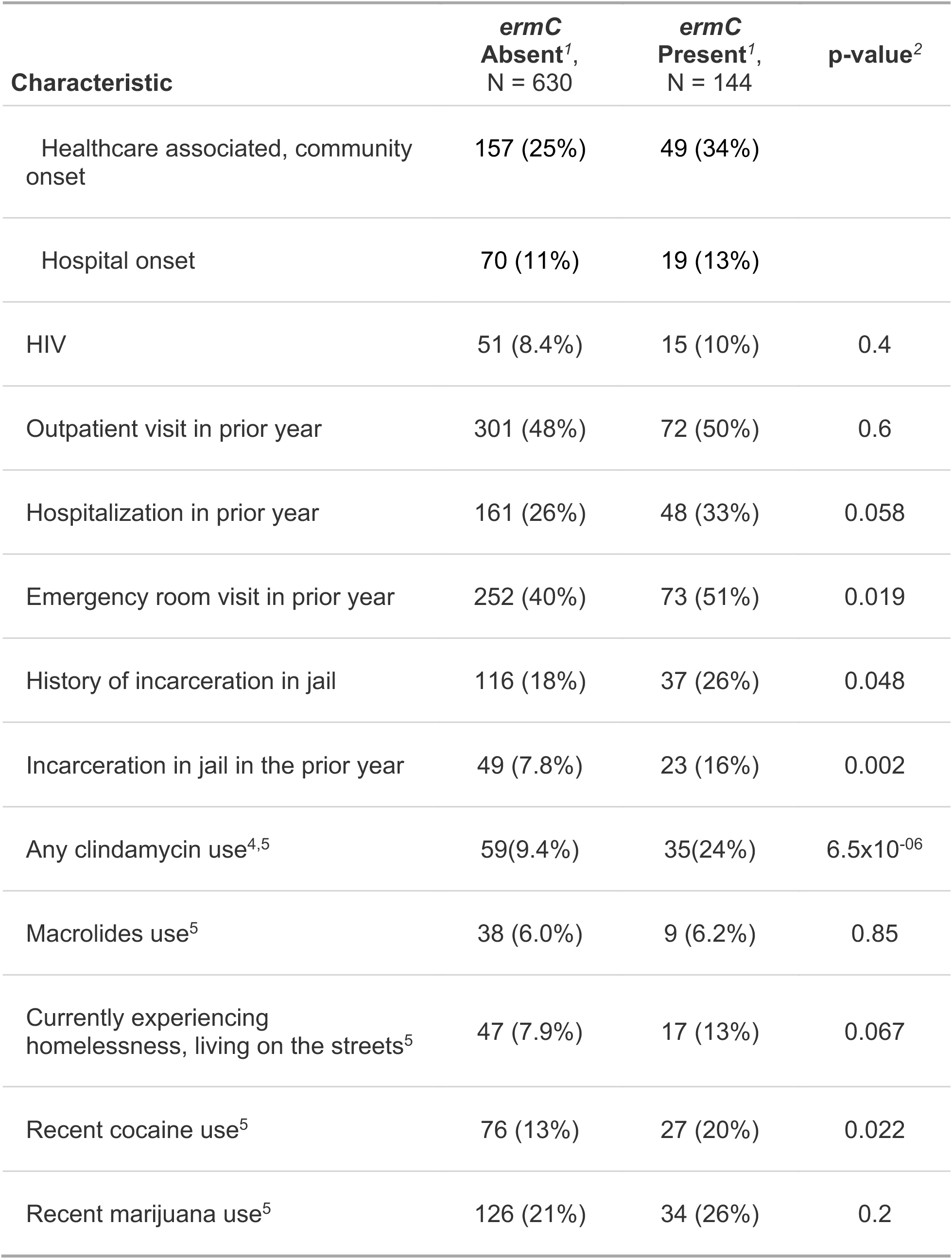
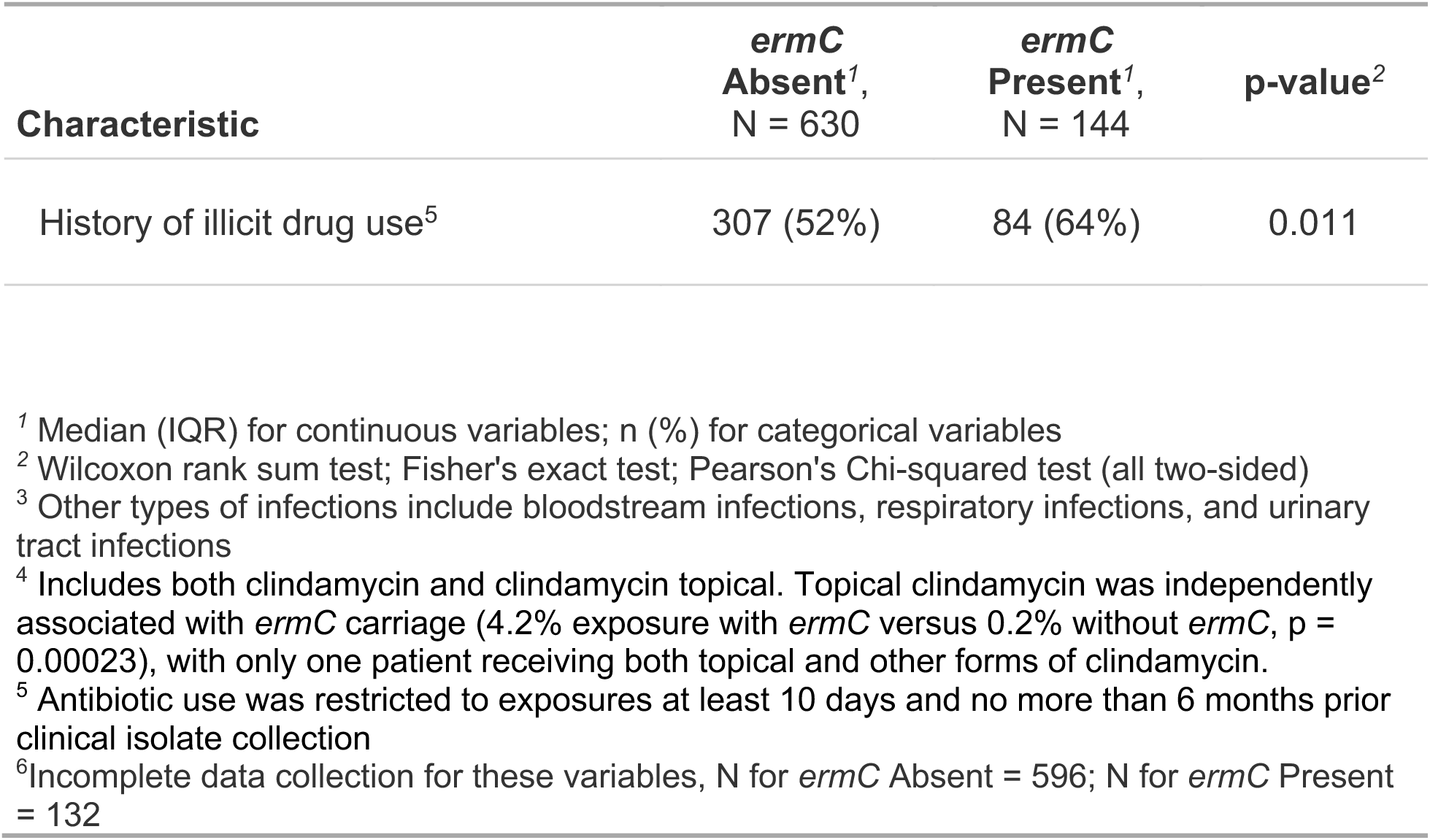
Epidemiologic factors associated with *ermC* plasmid in CCH cohort.

| Characteristic | *ermC*<br>Absent<sup>1</sup>,<br>N = 630 | *ermC*<br>Present<sup>1</sup>,<br>N = 144 | p-value<sup>2</sup> |
| --- | --- | --- | --- |
| Age | 43 (28, 52) | 44 (32, 53) | 0.3 |
| Race/ethnicity |  |  | 0.2 |
| Hispanic/Latino | 144 (23%) | 27 (19%) |  |
| Non-Hispanic Black | 370 (59%) | 89 (62%) |  |
| Non-Hispanic White | 102 (16%) | 21 (15%) |  |
| Other/Unable to determine | 14 (2.2%) | 7 (4.9%) |  |
| Sex |  |  | 0.2 |
| Female | 173 (27%) | 48 (33%) |  |
| Male | 457 (73%) | 96 (67%) |  |
| Wound infection<sup>3</sup> | 529 (84%) | 129 (90%) | 0.089 |
| Epidemiologic Categorization |  |  | 0.039 |
| Community onset | 403 (64%) | 76 (53%) |  |

| **Characteristic** | ***ermC***<br>**Absent<sup>1</sup>,**<br>**N = 630** | ***ermC***<br>**Present<sup>1</sup>,**<br>**N = 144** | **p-value<sup>2</sup>** |
| --- | --- | --- | --- |
| Healthcare associated, community onset | 157 (25%) | 49 (34%) |  |
| Hospital onset | 70 (11%) | 19 (13%) |  |
| HIV | 51 (8.4%) | 15 (10%) | 0.4 |
| Outpatient visit in prior year | 301 (48%) | 72 (50%) | 0.6 |
| Hospitalization in prior year | 161 (26%) | 48 (33%) | 0.058 |
| Emergency room visit in prior year | 252 (40%) | 73 (51%) | 0.019 |
| History of incarceration in jail | 116 (18%) | 37 (26%) | 0.048 |
| Incarceration in jail in the prior year | 49 (7.8%) | 23 (16%) | 0.002 |
| Any clindamycin use<sup>4,5</sup> | 59(9.4%) | 35(24%) | 6.5x10<sup>-06</sup> |
| Macrolides use<sup>5</sup> | 38 (6.0%) | 9 (6.2%) | 0.85 |
| Currently experiencing homelessness, living on the streets<sup>5</sup> | 47 (7.9%) | 17 (13%) | 0.067 |
| Recent cocaine use<sup>5</sup> | 76 (13%) | 27 (20%) | 0.022 |
| Recent marijuana use<sup>5</sup> | 126 (21%) | 34 (26%) | 0.2 |
| History of illicit drug use<sup>5</sup> | 307 (52%) | 84 (64%) | 0.011 |

<sup>1</sup> Median (IQR) for continuous variables; n (%) for categorical variables
<sup>2</sup> Wilcoxon rank sum test; Fisher's exact test; Pearson's Chi-squared test (all two-sided)
<sup>3</sup> Other types of infections include bloodstream infections, respiratory infections, and urinary tract infections
<sup>4</sup> Includes both clindamycin and clindamycin topical. Topical clindamycin was independently associated with *ermC* carriage (4.2% exposure with *ermC* versus 0.2% without *ermC*, p = 0.00023), with only one patient receiving both topical and other forms of clindamycin.
<sup>5</sup> Antibiotic use was restricted to exposures at least 10 days and no more than 6 months prior clinical isolate collection
<sup>6</sup> Incomplete data collection for these variables, N for *ermC* Absent = 596; N for *ermC* Present = 132

Having observed that clindamycin use was associated with *ermC* in both CCH and CCJ, we next compared frequency of clindamycin use between the two settings to assess the relative strength of the selective pressure imposed. This comparison revealed that clindamycin exposure was significantly higher among detainees in CCJ than patients in CCH, for both overall (21.2% vs. 12.1%, p = 0.033, **Figure 3B**) and topical use (7.5% vs. 0.90%, p = 1.1×10^−7^, **Figure 3B**). This led us to hypothesize that high rates of clindamycin use among detainee populations, beyond just those with detected MRSA colonization or infection in our study population, could be driving spread of *ermC*. In support of this hypothesis, we found more individuals in CCJ were prescribed topical clindamycin (range: 60-310 individuals per month) than were infected with MRSA (range: 0-12 per month), in any given month of our study period. Moreover, the use of topical clindamycin spiked in the CCJ just as our collection of MRSA in CCJ commenced in 2016 (**Supplemental Figure S7**).

### Genomic and epidemiologic evidence supports ermC amplification in the jail spilling over into the community

Having observed evidence for amplification of *ermC* in both CCJ and CCH, we next evaluated evidence for transmission in the jail influencing *ermC* prevalence in the community. First, we examined whether prior exposure to jail increased the risk of an individual’s isolate harboring *ermC*. Indeed, we found that among individuals at CCH, incarceration within the year prior to their MRSA clinical culture was associated with their strain harboring *ermC* (OR = 2.2, p = 0.004, 95% CI: 1.3-3.9). Other than clindamycin, healthcare, and jail exposures, the only other epidemiologic factor significantly associated with having a strain harboring *ermC* was current cocaine use or history of illicit drug use (**Table 1**). Of note, cocaine use and recent incarceration are associated (OR = 2.2, p = 0.01, 95% CI: 1.2-4.0), so it is possible that incarceration may be mediating or confounding the association between cocaine/illicit drug use and *ermC*. History of illicit drug use was also a risk factor for *ermC* harboring MRSA strains on intake surveillance at the jail, which likely reflects community transmission pre-detention and corroborates the associations found in the CCH data (**Supplemental Table S5**).

To gain further insights into the relationship between *ermC* and exposure to the CCJ, we took advantage of additional isolates collected from CCH from 2004-2020. While these additional isolates outside of 2011-2014 are less comprehensively sampled (i.e. only random subsets of wound isolates sequenced from 2004-2009 and only MRSA bloodstream infection isolates available 2015-2018), they allowed us to evaluate *ermC* prevalence and its association with recent exposure to jail over a longer timeframe. We observed a higher prevalence of *erm*C in individuals with recent incarceration across all study years (Χ^2^ = 9.8, p = 0.002) (**Figure 4A**). In addition, we observe an increasing prevalence of *erm*C overall (Χ^2^ test for trend from 2004-2015 = 38.3, p < 0.001), until a plateau in 2019-2020. As noted above, clindamycin use in the jail spiked in 2016, and then decreased to a low level in 2018 that persisted onward (**Supplemental Figure S7**).

**Figure 4:**
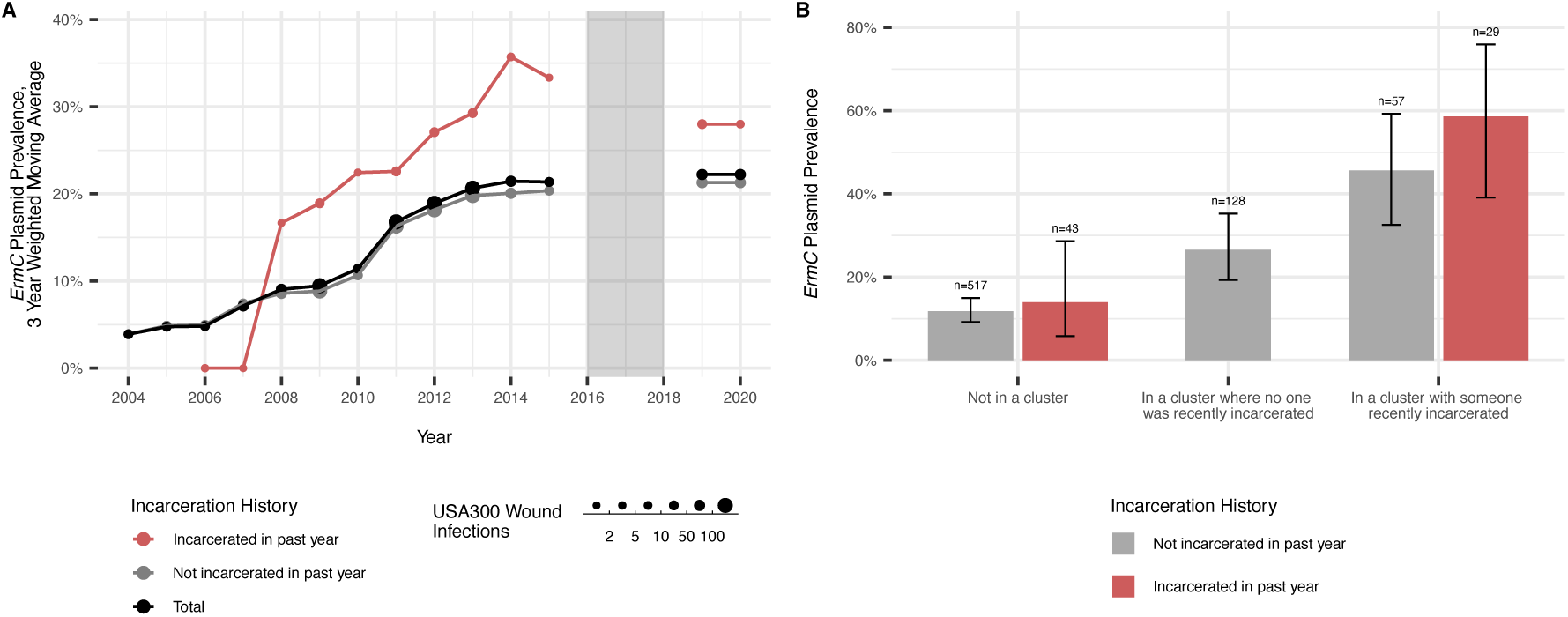
Relationship between incarceration history and *ermC* in the community. **A)** Prevalence of *ermC* in USA300 clinical wound isolates collected by CCH from 2004-2020. Values are smoothed and represent centered 3 year moving averages (or 2-year averages if data from an adjacent year were unavailable) weighted by number of observed infections per year. Note that from 2016-2018 (grey box) only bloodstream infection isolates were collected, so these years are not included in the figure. The red line represents prevalence of *ermC* in wound infections of those who have been incarcerated in the last year, the grey line represents infections of community members who had not been incarcerated in the last year, and the black line is total prevalence of *ermC* over time. **B)** The presence of *ermC* was determined for all MRSA isolates from CCH from 2011-2014. We observed that isolates in genomic clusters (within 20 SNVs of other isolates) were more likely to have *ermC* present than isolates not in genomic clusters, and this association was even greater in genomic clusters that included individuals who had been recently incarcerated (in the year prior to MRSA infection). When controlling for type of cluster membership, individual incarceration history (incarcerated in past year: red, not incarcerated in past year: grey) was not an additional predictor of *ermC* presence. That is, *ermC* prevalence was similar in those in a cluster with someone who was recently incarcerated whether that individual was recently incarcerated or not. Those who were recently incarcerated but not in a genomic cluster had similarly low prevalence of *ermC* as unclustered individuals who were not recently incarcerated. Error bars represent the 95% confidence interval for proportions and the numbers above the error bars represent sample size for each group.

Lastly, having seen evidence that direct exposure to the jail was associated with having an *ermC* carrying strain, we next set out to see whether indirect exposure to the jail, as evidenced by a genomic linkage of a MRSA isolate from individuals without history of incarceration to a MRSA isolate from someone with exposure to the jail, increased risk for having an *ermC* carrying strain. Indeed, we found that individuals in a genomic cluster with someone recently incarcerated have 6.7 times increased odds (95% CI: 4.0-11.3) of having *ermC* as those not in a genomic cluster, controlling for individual recent incarceration history (**Figure 4B**). Additionally, those who were in a genomic cluster that did not include someone who was recently incarcerated had 2.7 times increased odds (95% CI: 1.7-4.4) of having an *ermC* isolate compared to those who were not in a genomic cluster, further supporting the evidence that regardless of jail exposure (direct or indirect), *ermC*-carrying isolates may be associated with greater transmission in the community (**Figure 4B**).

## Discussion

It has been hypothesized that urban jails and prisons act as key amplifiers of CA-MRSA spread(17). However, in practice it has been challenging to separate the impact of transmission in jails and prisons from other socioeconomic community risk factors that are correlated with high detainee release. Here, through sampling circulating MRSA strains from a large urban jail and a healthcare network serving the surrounding communities, employing GWAS to identify variants preferentially spreading in jail settings, and leveraging detailed epidemiologic metadata, we find evidence of amplification of an antibiotic resistance element in MRSA isolates in the jail with re-seeding back into the surrounding communities. More broadly, this work demonstrates how genomic analysis of bacterial pathogens can yield not just insight into transmission pathways, but when combined with relevant metadata, also provide insight into the locations and practices mediating the emergence and spread of genotypes of concern.

Previous studies have shown that communities with high detainee release have elevated MRSA infection rates(14, 15), thereby suggesting a role for jails in the amplification of MRSA in connected communities. We previously found support for MRSA spread in an urban jail in the form of genomic and epidemiologic evidence of MRSA transmission among detainees(20). Here, we built on this finding and showed that jails also have the potential to act as amplifiers of specific genetic variants, in this case, the mobilizable clindamycin resistance-conferring *ermC* plasmid. Support for *ermC* amplification in the jail comes from its strong association with intra-jail genomic transmission linkages, as well as the 4-fold increase in *ermC* prevalence among jail-acquired MRSA isolates relative to isolates collected on entrance to the jail. Of note, the amplification was not due to the clonal spread of a single lineage, but rather the simultaneous spread of multiple USA300 sub-lineages that independently acquired the *ermC* plasmid. Evidence of downstream impact on the community comes from finding that the presence of *ermC* in community MRSA isolates is associated with recent individual exposure to the jail and with genetic linkage to isolates collected from individuals with exposure to the jail. Moreover, the plateau in the *ermC* prevalence in the years following a decrease in the use of topical clindamycin in the jail is consistent with a role of antibiotic use among detainees in amplifying antibiotic resistance in the jail and connected communities.

We believe our findings support the need for antibiotic stewardship efforts to extend beyond healthcare settings in locations such as correctional facilities. The association of prior topical clindamycin exposure with *ermC* carriage (12% with *ermC* versus 1.5% without *ermC*), combined with the significantly increased exposure to topical clindamycin in the jail versus community (7.5% versus 0.90%), supports the role for antibiotic use in the jail playing a role in amplifying *ermC*, and more broadly showing how antibiotic usage in individual settings can have broader impact on antibiotic resistance. As another example of the distributed impact of antibiotic use, it is increasingly appreciated that long-term care settings and nursing homes that have high rates of antibiotic use and exchange large numbers of patients with regional healthcare facilities, can have a significant impact on regional prevalence of antibiotic resistance threats(27–29). Together, these observations raise the possibility of targeting infection prevention and antibiotic stewardship interventions to these key community and healthcare hub facilities to maximize the impact of finite resources available for regional control of antibiotic resistance.

From an analytic perspective, our work demonstrates the potential for regional genomic epidemiology of bacterial pathogens to track the emergence of variants of concern and elucidate their drivers. Complexity in modes of bacterial genome evolution have to this point hindered the direct application of phylodynamic approaches that have been successfully employed in viral pathogens to detect variants of concern(30). However, work in the clonal bacterial pathogen *Mycobacterium tuberculosis* has shown the promise of phylodynamic approaches in bacteria, revealing preferential amplification of drug resistant and susceptible lineages in specific host populations(31, 32). Here we took an unbiased GWAS approach to identify not just lineages, but rather variants associated with recent amplification, leading to the detection of *ermC* as preferentially spreading in both CCJ and CCH collections. Of note is the complex distribution of *ermC*, where its amplification was not due to the expansion of a single sub-lineage, but rather the expansion of multiple sub-lineages that had independently acquired the *ermC* plasmid. This observation highlights the need to consider nuances of bacterial evolution in future studies leveraging methodologic innovations from viral genomic analysis. Lastly while we had unique access to samples from CCH and CCJ, our sampling primarily consisted of clinical cultures, demonstrating the feasibility of assembling collections needed to power these analyses and thereby highlighting the promise of regional genomic surveillance.

Our study has some limitations that should be considered when interpreting our findings. First, while we had access to comprehensive collections of clinical isolates from both CCJ and CCH, the collection periods were not overlapping (CCH: 2011-2014 and CCJ: 2015-2018). However, despite not being able to look at direct transmission via comparison between isolates collected from CCJ and CCH, we were able to use knowledge of recent jail exposure among individuals seeking care at CCH to link the jail and community. A second limitation of our study is that the majority of isolates in our collection represent clinical infection, with the CCH collection in particular only including clinical isolates. However, inclusion of MRSA colonization isolates from detainees at jail intake support high prevalence of *ermC* in the community, and our conclusions regarding *ermC* amplification are supported by the steady longitudinal increase in *ermC* prevalence at CCH.

In conclusion, through the integration of genomic and epidemiologic data from an urban jail and the connected community, we find evidence supporting the preferential spread of clindamycin-resistant MRSA in the jail, with spread into the community. Moreover, integration of antibiotic usage data highlighted the role of clindamycin use in both jail and community settings in selecting for the horizontal gene transfer of the *ermC*-carrying plasmid and spread of strains already harboring the plasmid. In addition to immediate implications for antibiotic stewardship, this work highlights more broadly the potential for genomic surveillance to reveal the sources and drivers of emerging infectious threats in the community. However, essential to generating translational insights capable of guiding intervention is the careful sampling of key community reservoirs, as well as collection of relevant epidemiologic data that enhances understanding of pathways of transmission and their clinical, societal, and behavioral amplifiers.

## Methods

### USA300 MRSA isolate genomes and metadata from Cook County Jail and Cook County Health

The current and previous studies from which data were used were approved with waiver of consent by the CCH Institutional Review Board. Metadata and MRSA sequences from two previous studies, as well as a new collection, were analyzed here (See **Supplementary Tables S1 and S6** for comparison of populations and **Table S7** for summary of included genomes). The first dataset captures all clinical cultures presenting to Cook County Health (CCH) from 2011-2014 (Bioproject PRJNA734638)(25). CCH is the major healthcare network in Chicago, Illinois. We previously showed evidence of community acquisition of those presenting with MRSA infection to CCH(25), leading us to consider samples collected at CCH as a representation of MRSA USA300 strains circulating in the community. The second data set includes surveillance isolates for nasal and extra-nasal MRSA colonization at jail intake and at day 30 for a subset of detainees from 2015-2017, as well as all MRSA isolates derived from detainee clinical infections during the same time period (Bioprojects PRJNA638400, PRJNA530184, and PRJNA761409)(18–20). In addition to previously published data sets, we sequenced additional genomes from CCH spanning 2004-2020 to expand our ability to examine trends in MRSA over time (Bioproject PRJNA1225712). The newly sequenced genomes were selected to randomly capture circulating USA300 over time by preferentially selecting for sequencing clinical isolates cultured from skin and soft-tissue infections. The exception to this was for the years 2015-2018, when only bloodstream infection isolates were available.

### Genome sequencing and analysis

New genomes from CCH were sequenced on Illumina Novaseq instruments at the Advanced Genomics Core at the University of Michigan. The quality of sequencing reads was assessed using FastQC v0.11.0, and adapter sequences and low-quality bases removed using Trimmomatic v0.39. SNVs were identified by first using Burrows-Wheeler short-read aligner (bwa v0.7.17) to map trimmed reads to the USA300 reference genome (GenBank accession number NC_010079.1), then discarding polymerase chain reaction (PCR) duplicates with Picard v3.0.0, and calling variants with SAMtools and bcftools v1.9. Variants were filtered using VariantFiltration from GATK v4.5.0.0 (QUAL > 100; MQ>50;>=10 reads supporting variant; and FQ< 0.025). We performed GATK HaplotypeCaller for indel calling only including those with root mean square quality (MQ) > 50.0, GATK QualbyDepth (QD) > 2.0, read depth (DP) > 9.0, and allele frequency (AF) > 0.9. We also excluded variants that were less than 5 base pairs in the proximity to indels, in recombinant regions identified by Gubbins v3.0.0, in a phage region identified by Phaster web tool, or resided in tandem repeats of length greater than 20 bp as determined using the exact-tandem program in MuMmer v3.23 using a custom Python script. We conducted multilocus sequence typing with ARIBA and further classified isolates using in silico sequencing probes provided in Bowers et al. (33, 34).

### Phylogenetic analysis

The recombination-masked whole-genome alignment was then used to reconstruct a maximum likelihood phylogeny with IQ-TREE v2.0.3 using the general time reversible model GTR+G and ultrafast bootstrap with 1000 replicates (-bb 1000).

### Testing the association of potential transmission events and the MRSA pangenome

We used panaroo v 1.2.5 (clean mode moderate) to identify the accessory genome of each sample(35). Genes were annotated with eggnog v2.1.12(36), with the following parameters: -m diamond --override --itype CDS --translate -- report_orthologs. Individuals linked by recent direct or indirect transmission were defined with a 20 SNV threshold, based on prior literature identifying optimal thresholds for *S. aureus*(21, 37), and supported by our previous analysis of these particular datasets(20, 25). A Fisher’s exact test was conducted to assess the association between transmission and gene using the R package exact2×2 v1.6.5(38). The relationship between odds ratio (OR) and p value was plotted.. Significance was assessed with a Bonferonni-adjusted p value. We made the intentional decision not to explicitly control for population structure in our GWAS, as the clustering of strains is the signal we were trying to identify a genetic basis for. For example, using regression-based approaches that account for population structure by including a distance-matrix as a random effect, would control for the genetic clustering we are trying to detect(39). In contrast, tools that identify lineages that are undergoing clonal expansions, do not currently capture the simultaneous expansion of multiple lineages due to mobile genetic elements(40). However, not controlling for population structure opens the analysis to simply detecting lineage effects, which may or may not be causal. It is for this reason that we specifically focus on the accessory genome, which by way of mobile elements mediating horizontal spread, break the linkage with genetic background.

### Ancestral state reconstruction to evaluate the emergence and spread of *ermC* plasmid-containing strains

The emergence and spread of *ermC* plasmid-containing strains was characterized using the R package *phyloAMR* (https://github.com/kylegontjes/phyloAMR)(41). *PhyloAMR*’s *asr()* function applied corHMM’s joint ancestral state reconstruction on the *ermC* plasmid and traced ancestral- and tip-states across edges of the midpoint-rooted phylogeny, comprising both CCH and CCJ isolates, to infer gain events and loss events(42). The all-rates different model was determined as the best rate matrix using sample-size corrected Akaike information criterion.

The phylogenetic tree-traversal algorithm, *asr_cluster_detection()*, inferred the evolutionary history of *ermC* in this population. The phylogeny was traversed from tip to root to classify *ermC* plasmid-containing isolates as phylogenetic singletons (i.e., evidence of independent acquisitions of the plasmid) or members of a phylogenetic cluster of *ermC* plasmid-containing strains (i.e., evidence of the emergence and spread of a plasmid-containing lineage). This algorithm classified *ermC* plasmid-containing isolates with gain events at their tip as phylogenetic singletons. However, isolates with a gain event at the tip and a loss event at their parental node were eligible for classification as members of a phylogenetic cluster. Isolates with ancestral gain events that were shared with at least one additional *ermC*-containing isolate contributed to the study by another individual were classified as members of a phylogenetic cluster. Isolates that did not share an ancestral gain event with another *ermC* plasmid-containing isolate were classified as phylogenetic singletons.

### Antibiotic susceptibility testing and genetic determinants of clindamycin resistance

Antibiotic susceptibility testing was performed on all clinical isolates (i.e. not for surveillance isolates) in the Cook County Health clinical microbiology laboratory using Microscan susceptibility testing automated system, with resistance determined using CLSI breakpoints. The presence of *ermC* can confer constitutive or inducible resistance to clindamycin(24). We defined inducible resistance as presence of *ermC,* along with susceptibility to clindamycin and resistance to erythromycin by broth microdilution; we defined constitutive resistance as presence of *ermC,* along with resistance to both erythromycin and clindamycin. Furthermore, we used pyseer(39) to confirm that the *ermC*-carrying plasmid was the only determinant of clindamycin resistance in this collection (**Supplemental Figure S8)**.

### Epidemiological associations with presence of the *erm*C-carrying plasmid

The presence of the *ermC*-carrying plasmid was determined by mapping reads using bwa v0.7.17 to the USA300-SUR4 reference plasmid (GenBank accession number NZ_CP014374.1). The plasmid was determined to be present in a given genome if the mean mapping coverage to the SUR4 plasmid was greater than the mean mapping coverage to the USA300 reference chromosome (GenBank accession number NC_010079.1). We assessed the association between binary epidemiological factors and presence of the *erm*C-carrying plasmid with a Fisher’s exact testing using the R package exact2×2. We examined previously collected demographic data, comorbidities, prior healthcare exposures, unstable housing, substance abuse, and history of incarceration(19, 20, 25). We used administrative pharmacy data at the CCJ to evaluate topical clindamycin use (i.e., soaked cloths and solution) among detainees who sought medical care. Topical clindamycin was primarily prescribed for acne and other dermatology related conditions, although our data pull was in aggregate so we cannot rule out that there were additional indications for its use. We used Poisson regression to determine the association between cluster size and rate of *ermC* prevalence within clusters in both the CCJ and community datasets. We used logistic regression to explore the association between type of cluster (unlinked vs cluster with no one incarcerated in the past 6 months vs cluster with someone incarcerated in the past 6 months), and isolate *ermC* presence, while controlling for individual recent incarceration history.

## Data availability

Whole-genome sequences analyzed in this study have been uploaded to the SRA under Bioprojects PRJNA734638, PRJNA638400, PRJNA530184, PRJNA761409 and PRJNA1225712. The raw individual level meta-data are protected and are not available due to data privacy laws, but aggregate population summaries statistics are available in main text and supplementary tables.

## Code availability

Select analysis code available at https://github.com/sthiede/thiede_steinberg_et_al_MRSA_ermC/ (DOI: 10.5281/zenodo.17350441).

## Funding and acknowledgments

The project described was supported by Grant Numbers R01AI114688 (PI: KJP) and 1R01 AI146079-01A1 (PI: KJP) from the National Institute of Allergy and Infectious Diseases. SNT was supported by the Molecular Mechanisms of Microbial Pathogenesis training grant (NIH T32 AI007528). KJG was supported training fellowships from National Human Genome Research Institute (T32-HG000040) and NIAID (F31-AI186288). ESS was supported by NIAID U19AI181767. We would also like to thank Joshua Rafinski and Will Chan for their assistance with data analysis. We thank the individuals who participated in this study.

## Author Contributions

SNT, ESS and KJP conceived of the study. AA, NMM, CZ and SS processed samples and clinical data. SNT, HS, EB, KJG, KW, SR and AP performed data analysis. SNT, HS, RAW, KJP and ESS drafted the manuscript.

