## Supplemental Tables and Figures for "Antibiotic-resistance plasmid amplified among MRSA cases in an urban jail and its connected communities"

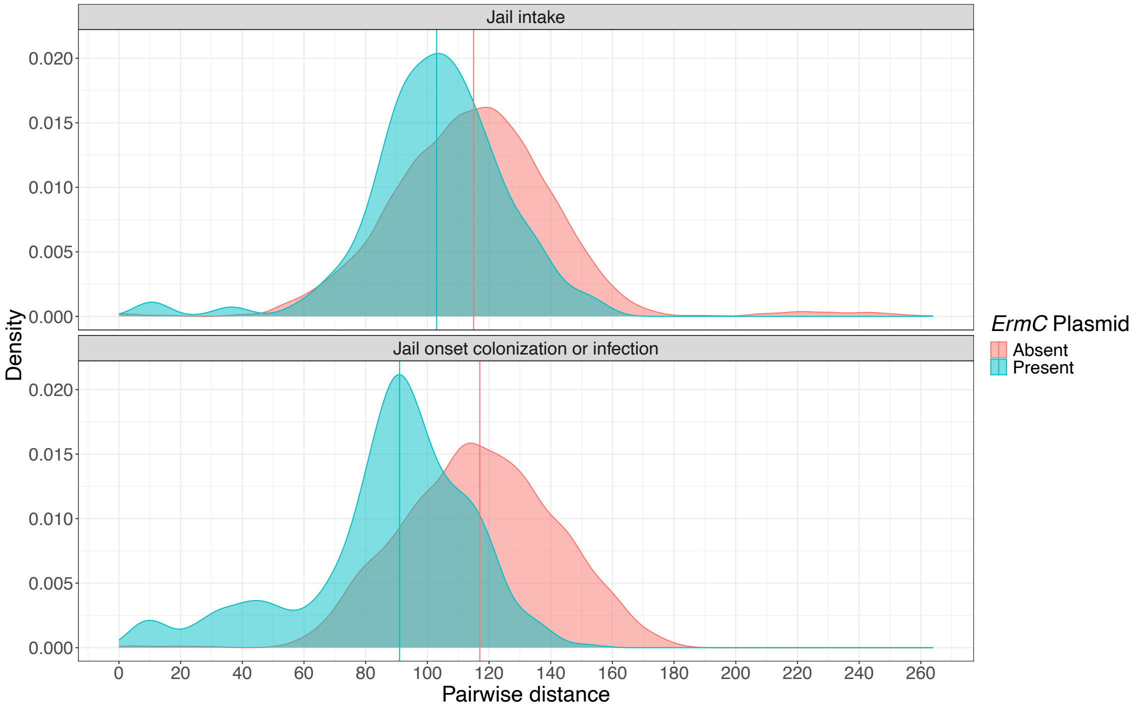


**Supplemental Figure S1: Recent expansion of ermC in the jail.** Pairwise distances of USA300 MRSA isolates with *ermC* (teal) are shorter on average than those without *ermC* (salmon), and this especially true in jail onset colonizations or infections (bottom) compared to jail intake cases (top). This suggests that *ermC*-carrying isolates in the jail may be trasmitted more frequently than non-*ermC* isolates. Vertical lines represent median pairwise distances for each group.

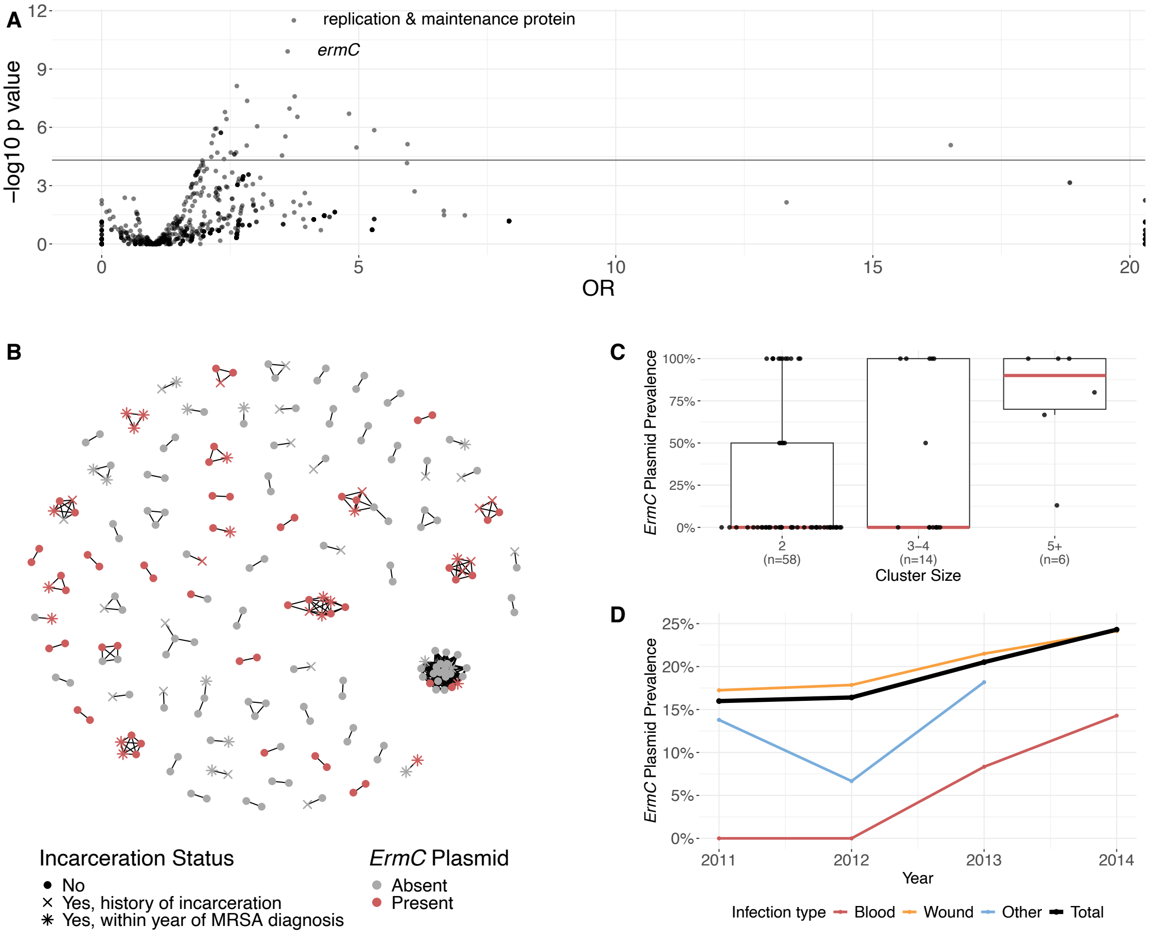


**Supplemental Figure S2: Association between pangenome and transmission in the community, 2011-2014. A)** A genome-wide association study was performed to measure the association between accessory gene presence and genetic linkage to another CCH isolate within 20 SNVs. P-values represent unadjusted p-values from two-sided Fisher's exact tests. Horizontal line indicates Bonferonni adjusted p-value threshold. **B)** Network diagram of MRSA transmission clusters. Edges indicate that isolates are related within 20 SNVs. Nodes are colored by presence (red) or absence (grey) of the *ermC*-carrying plasmid in the corresponding isolate. Shape indicates the isolate type, where circles are isolates from individuals with no incarceration history, exes are individuals with a history of incarceration (but not in the last year), and stars are those who were incarcerated in the year before MRSA diagnosis. There were 560 unclustered isolates in our analysis. Of these, 67 (12.0%) had the *ermC* plasmid present, 43 (7.7%) were incarcerated within the year before MRSA diagnosis, and 55 (9.8%) were incarcerated but more than a year before MRSA diagnosis. Of those unclustered isolates from individuals who did not have a history of incarceration, 56 (12.1%) had the *ermC* plasmid present, 6 (14.0%) of unclustered isolates in individuals with recent incarceration history had the *ermC* plasmid, and 5 (9.1%) of unclustered isolates in individuals with a history of incarceration more than one year before MRSA diagnosis had the *ermC* plasmid. **C)** Larger clusters of MRSA isolates are associated with higher *ermC* prevalence than small clusters. Boxes represent the interquartile range (IQR) of *ermC* plasmid prevalence, median *ermC* plasmid prevalence is represented by red horizontal lines, and whiskers represent the range of *ermC* plasmid prevalence in each cluster size group (clusters of 2 isolates, 3-4 isolates, or 5 or more isolates) excluding outliers (values more than 1.5 times the IQR from the 25th or 75th percentile for a given group). The number of clusters in each size category are indicated under the x-axis labels. **D)** Increasing prevalence of *ermC* among clinical MRSA cultures collected at CCH from 2011 to 2014. Red line represents blood infections, orange line represents wound infections, blue line represents other infections, and black line represents the increase in *ermC* plasmid prevalence overall.

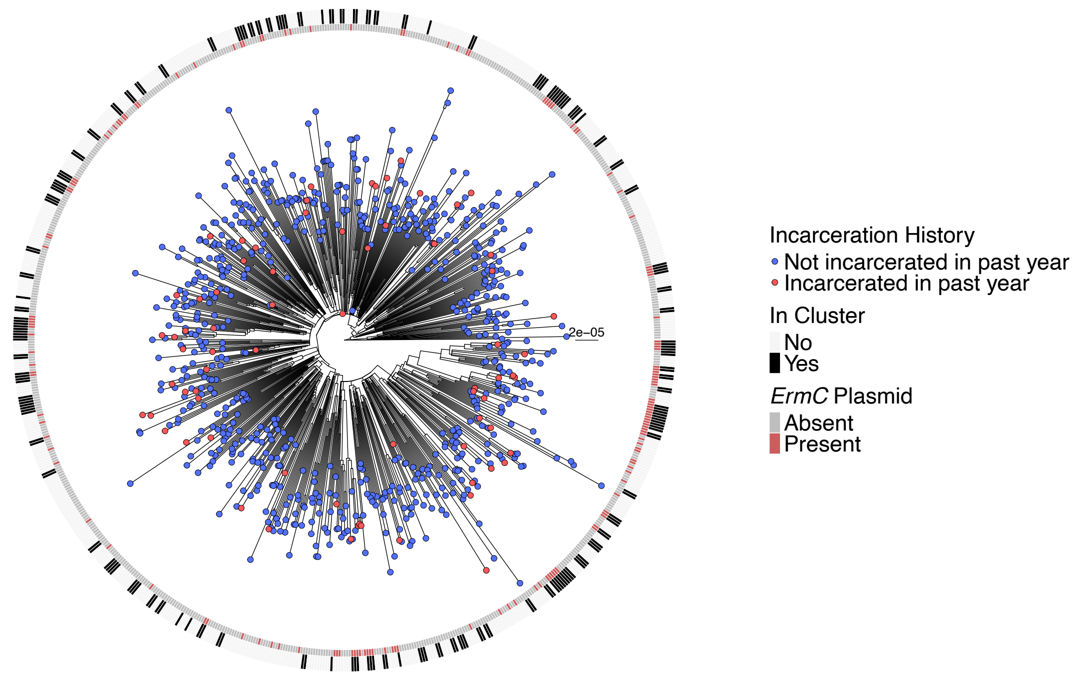


**Supplemental Figure S3: Phylogenetic tree of USA300 MRSA in the community, 2011-2014.** Maximum likelihood phylogeny of jail USA300 samples created with IQTREE using core-genome variants. Scale bar indicates substitutions per site. Tips designate if the patient was incarcerated in the last year (red) or not (blue). Inner ring indicates the presence (red) or absence (grey) of the *ermC*-carrying plasmid. Outer ring indicates if the isolate is genetically related to any other isolate within 20 SNVs (black) or not (white).

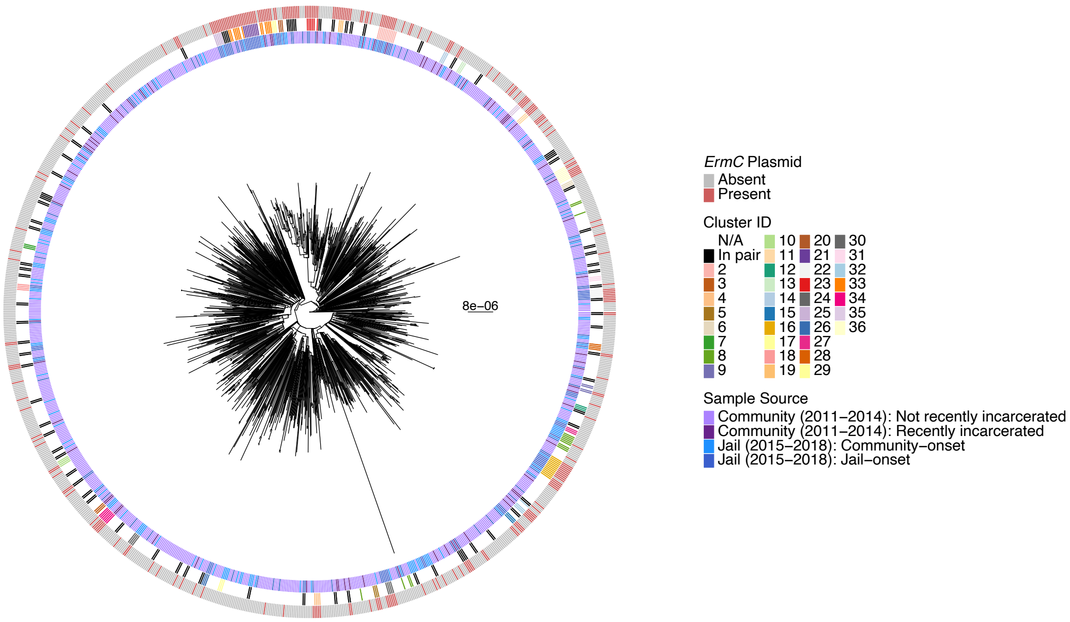


**Supplemental Figure S4. Phylogenetic tree of USA300 MRSA in the community (2011-2014) and the Cook County Jail (2015-2018).** Maximum likelihood phylogeny of jail USA300 samples created with IQTREE using core-genome variants. Scale bar indicates substitutions per site. Inner ring indicates the sample study source. Community infection isolates are shown in purple with isolates from recently incarcerated individuals (in the past year) shown in dark purple and others in light purple. Jail isolates are shown in blue with community-onset infections/intake colonizations represented by light blue and jail-onset infections/colonizations represented by dark blue. Middle ring indicates if the isolate is in a cluster with any other isolate. Isolates in a cluster consisting of just two isolates are represented in black, clusters with n > 2 are each represented with their own color, and unclustered isolates are represented in white. Outer ring indicates the presence (red) or absence (grey) of the *ermC*-carrying plasmid.

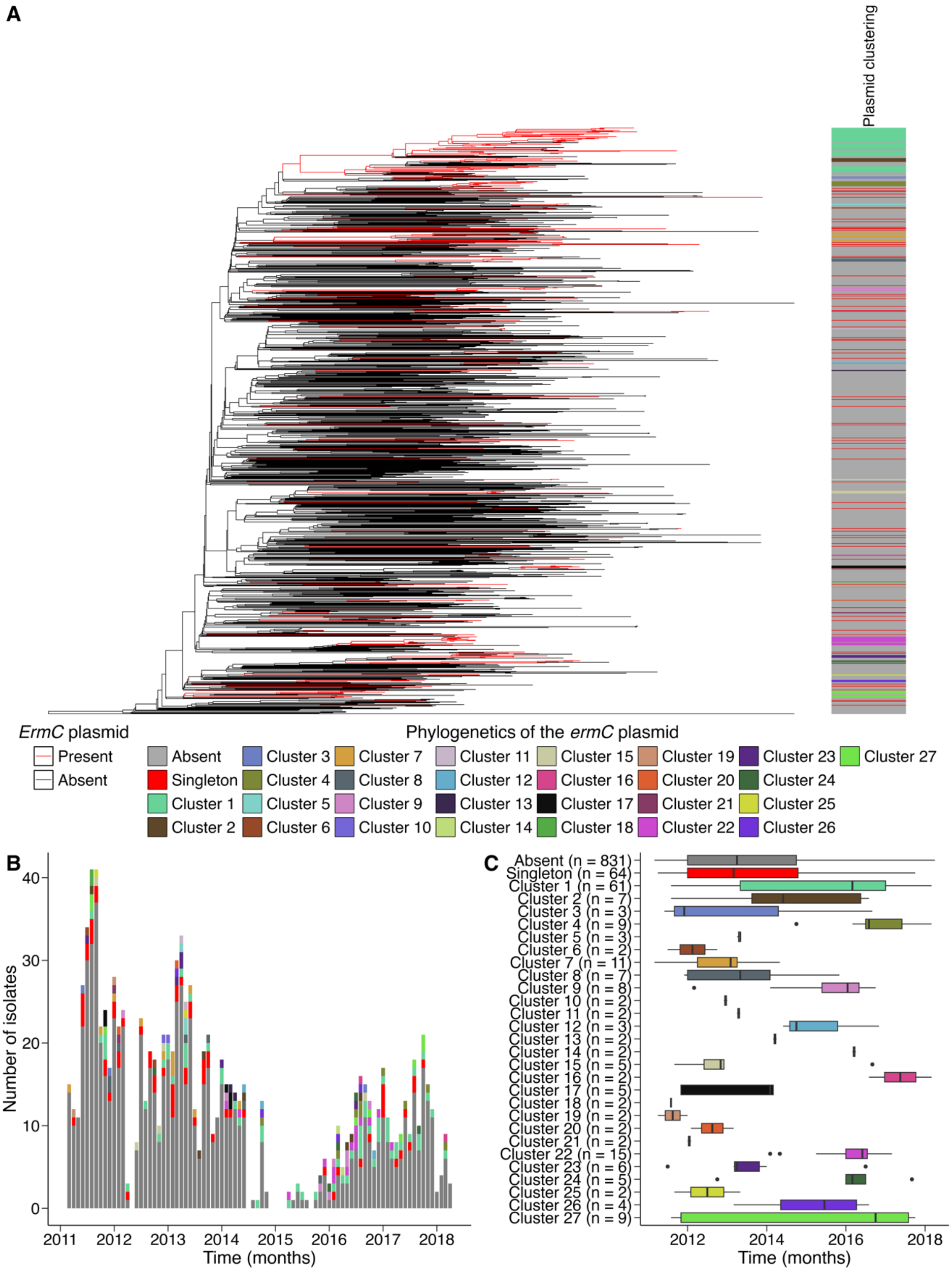


**Supplemental Figure S5. Frequent emergence and spread of *ermC* plasmid-containing strains. (A)** Clustering of the *ermC* plasmid across the midpoint-rooted USA300 phylogeny of Cook County Health and Cook County Jail isolates phylogenetic tree. Ancestral states, as inferred using joint ancestral state reconstruction under the all-rates-different model, were depicted across the edges of the phylogeny, with the presence of the plasmid indicated in red. Phylogenetic clustering was inferred by tracing from tip to root to classify *ermC* plasmid-containing isolates as phylogenetic singletons (i.e., evidence of independent acquisitions of the plasmid) or members of a phylogenetic cluster of *ermC* plasmid-containing strains (i.e., evidence of the emergence and spread of a plasmid-containing lineage). (**B)** Distribution of *ermC* plasmid-containing strains across the study period. (**C**) Duration of phylogenetic singletons and clusters in the study. Phylogenetic groups and the number of individuals in each group are indicated on the y-axis.

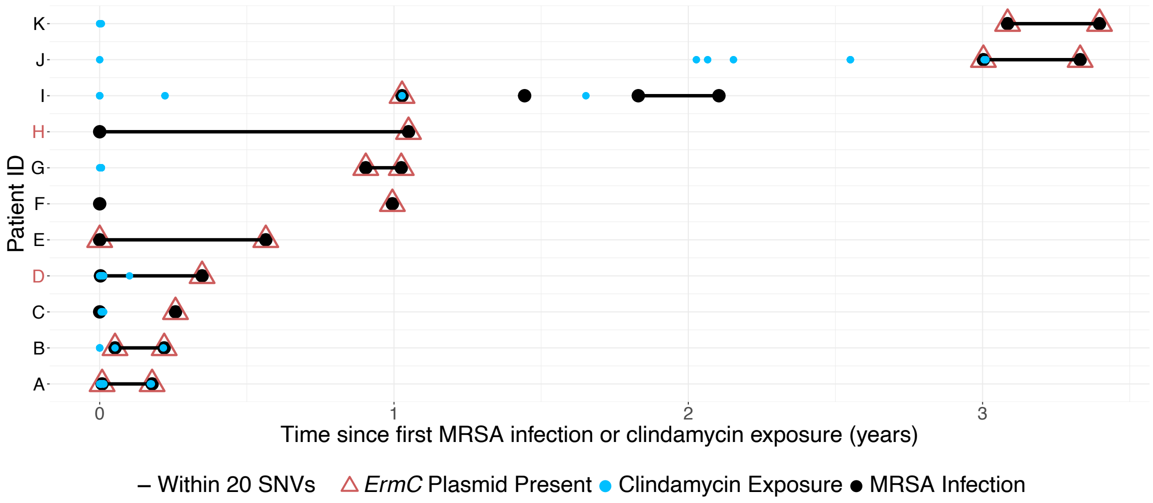


**Supplemental Figure S6: Acquisition of *ermC* within individual patients.** Focusing on patients from CCH with repeat clinical cultures (y-axis), we observe multiple instances of putative horizontal acquisition of the *ermC* plasmid in a non-plasmid carrying strain (red labels on y-axis). Red triangles indicate presence of *ermC*. Blue dots indicate clindamycin exposure. Black lines indicate infection of the same strain (within 20 single nucleotide variants, or SNVs, of the previous infection). Clindamycin exposure data before 2011 is unavailable. We observed 8 patients in the CCH dataset that had related isolates (within 20 SNVs) cultured at different times between 2011-2014, in which at least one of the pair of isolates had the *ermC* carrying plasmid. Of these pairs, there were two observations of an acquisition of the *ermC* plasmid,  6 observations of maintenance of the *ermC* plasmid, and no instances in which the first isolate carried the *ermC* plasmid and then the plasmid was lost by the time the second isolate was cultured. The median time between first and second cultures for the isolate pairs in which *ermC* was maintained was 88 days (range: 44-206 days) suggesting the plasmid can remain stable in the genome for extended periods of time.

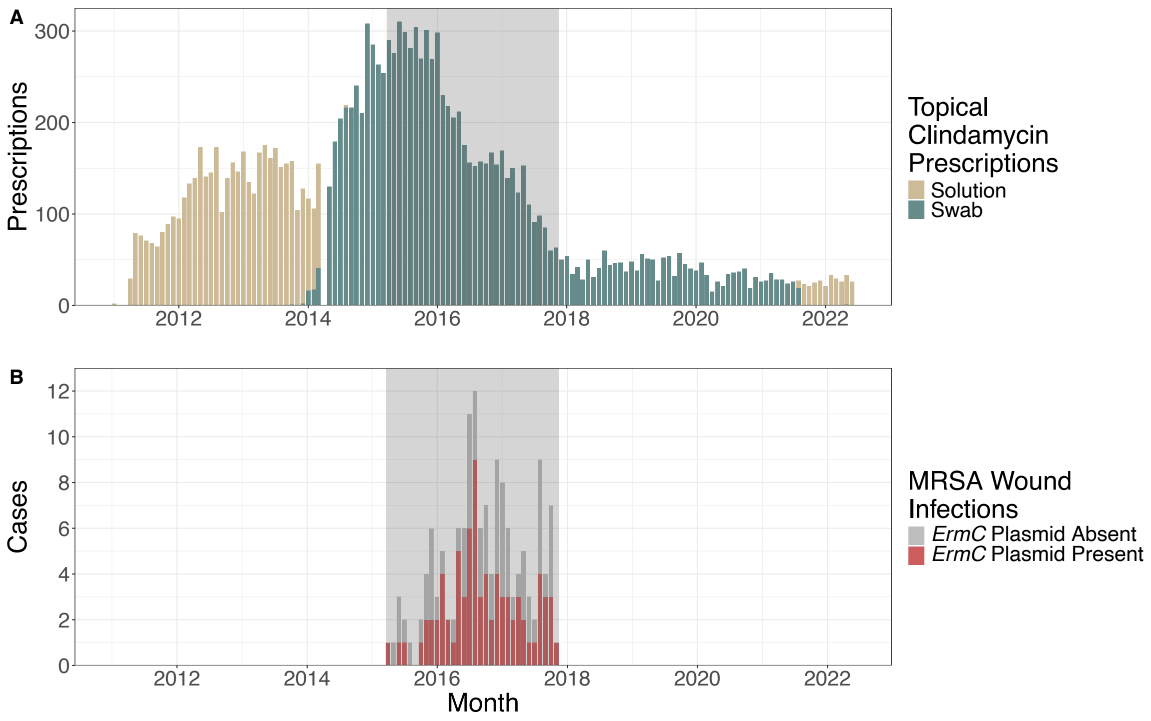


**Supplemental Figure S7: Clindamycin prescriptions and MRSA wound infections in the Cook County Jail. A)** Topical clindamycin prescriptions for all detainees at CCJ from January 2011 to June 2022. Topical clindamycin solution prescriptions are shown in tan and were most common from 2011-early 2014 and from late 2021-mid 2022. Topical clindamycin swab prescriptions are shown in teal and were most common from early 2014-late 2021. The grey box represents our study period; during this time, topical clindamycin swabs were exclusively prescribed, and rates of prescriptions dropped dramatically from roughly 300 prescriptions a month to less than 100. **B)** MRSA wound infections diagnosed in CCJ detainees during the study period (grey shaded box). *ErmC*-positive cases are shown in red and *ermC*-negative cases in grey. There is not a clear temporal association between the decrease in clindamycin prescriptions and total MRSA wound infections in the jail, although *ermC* positive cases do seem to decrease after prescriptions begin to decrease. Clindamycin prescriptions well outnumber MRSA infections throughout the study period, suggesting the topical medication was prescribed for conditions other than just MRSA infections.

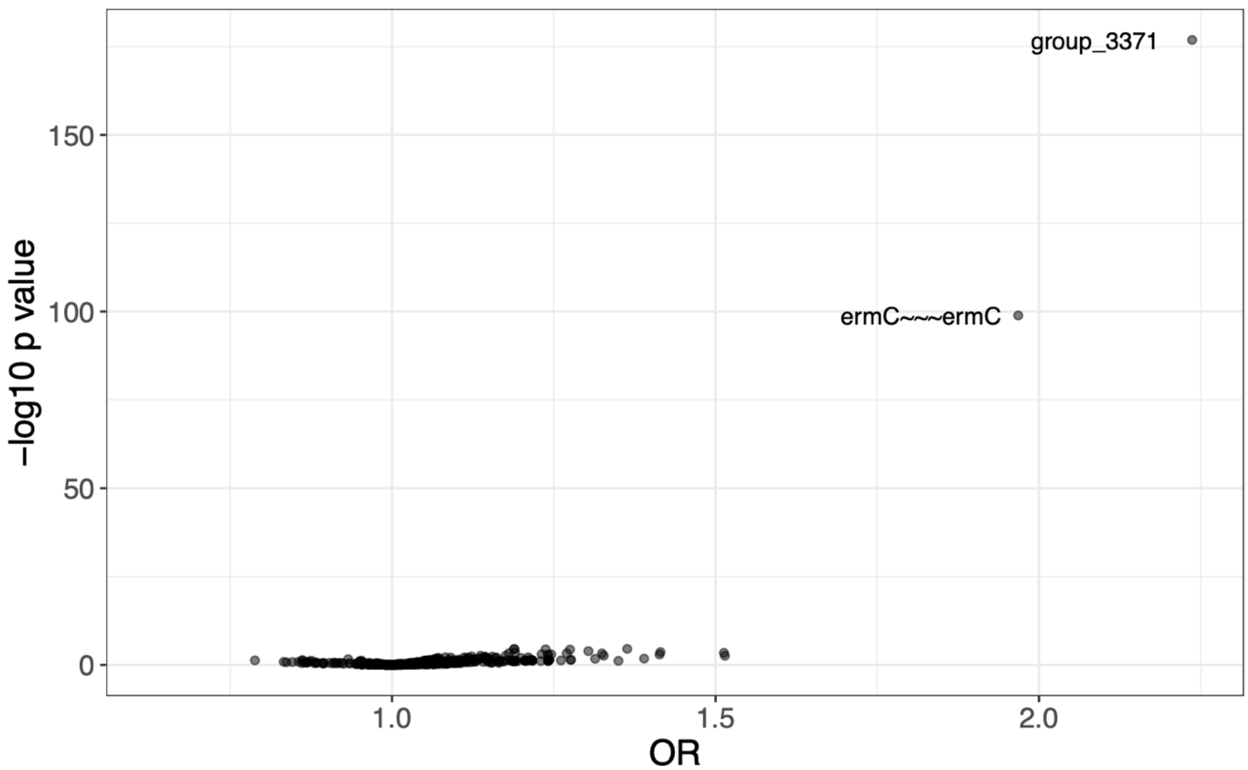


**Supplemental Figure S8:** Pyseer results assessing association between clindamycin resistance and the pangenome in the Cook County Health data (2011-2014). group_3371 indicates the replication and maintenance protein on the 2.4kb plasmid with *ermC*.

**Supplemental Table S1.** Demographic and clinical characteristics of MRSA colonized and infected detainees in the Cook County Jail (2015-2018).

| **Characteristic** | **N = 308**[1] |
|---|---|
| Year | |
| 2015 | 21 (6.8%) |
| 2016 | 118 (38%) |
| 2017 | 151 (49%) |
| 2018 | 18 (5.8%) |
| Sex | |
| Female | 79 (26%) |
| Male | 229 (74%) |
| Isolate type | |
| Community onset infection | 3 (1.0%) |
| Intake colonization | 147 (48%) |
| Jail onset colonization | 12 (3.9%) |
| Jail onset infection | 146 (47%) |
| Body site | |
| Abscess | 1 (0.3%) |
| Urine | 3 (1.0%) |
| Wound | 145 (47%) |
| Groin | 30 (9.7%) |
| Nose | 89 (29%) |
| Throat | 40 (13%) |
| *ErmC* plasmid present | 103 (33%) |

[1]n (%)

**Supplemental Table S2:** Presence of replication & maintenance gene and *ermC* determined by panaroo. Presence of USA300-SUR4 plasmid present if the mean mapping coverage to the SUR4 plasmid was greater than the mean mapping coverage to the USA300 reference chromosome. Here, 1 indicates presence and 0 indicates absence. Concordance between all 3 calls is shown in green shading; concordance is 98% (303/308) in CCJ and 94% in CCH (726/774).

CCJ:

| Presence of Replication & Maintenance Gene | Presence of ErmC Gene | Presence of Plasmid | n |
|---|---|---|---|
| 0 | 0 | 0 | 201 |
| 1 | 1 | 1 | 102 |
| 1 | 1 | 0 | 3 |
| 0 | 1 | 1 | 1 |
| 1 | 0 | 0 | 1 |

CCH:

| Presence of Replication & Maintenance Gene | Presence of ErmC Gene | Presence of Plasmid | n |
|---|---|---|---|
| 0 | 0 | 0 | 609 |
| 1 | 1 | 1 | 117 |
| 1 | 0 | 1 | 25 |
| 1 | 1 | 0 | 11 |
| 0 | 1 | 0 | 6 |
| 1 | 0 | 0 | 4 |
| 0 | 0 | 1 | 2 |

**Supplemental Table S3.** Characteristics of genomic clusters of MRSA isolates in the jail (2015-2017) and the community (2011-2014).

| | Cluster Size (number of isolates) | Number of isolates carrying *ermC* plasmid | Proportion of isolates carrying *ermC* plasmid | Location |
|---|---|---|---|---|
| 1 | 2 | 0 | 0.00 | Jail |
| 2 | 2 | 0 | 0.00 | Jail |
| 3 | 2 | 0 | 0.00 | Jail |
| 4 | 2 | 0 | 0.00 | Jail |
| 5 | 2 | 0 | 0.00 | Jail |
| 6 | 2 | 0 | 0.00 | Jail |
| 7 | 2 | 0 | 0.00 | Jail |
| 8 | 2 | 0 | 0.00 | Jail |
| 9 | 2 | 0 | 0.00 | Jail |
| 10 | 2 | 0 | 0.00 | Jail |
| 11 | 2 | 0 | 0.00 | Jail |
| 12 | 2 | 0 | 0.00 | Jail |
| 13 | 2 | 1 | 0.50 | Jail |
| 14 | 2 | 1 | 0.50 | Jail |
| 15 | 2 | 2 | 1.00 | Jail |
| 16 | 2 | 2 | 1.00 | Jail |
| 17 | 2 | 2 | 1.00 | Jail |
| 18 | 2 | 2 | 1.00 | Jail |
| 19 | 2 | 2 | 1.00 | Jail |
| 20 | 2 | 2 | 1.00 | Jail |
| 21 | 3 | 0 | 0.00 | Jail |
| 22 | 3 | 0 | 0.00 | Jail |
| 23 | 3 | 0 | 0.00 | Jail |
| 24 | 3 | 0 | 0.00 | Jail |
| 25 | 3 | 1 | 0.33 | Jail |
| 26 | 3 | 1 | 0.33 | Jail |
| 27 | 3 | 2 | 0.67 | Jail |
| 28 | 3 | 3 | 1.00 | Jail |
| 29 | 3 | 3 | 1.00 | Jail |
| 30 | 4 | 0 | 0.00 | Jail |
| 31 | 4 | 0 | 0.00 | Jail |
| 32 | 4 | 2 | 0.50 | Jail |

| | | | | |
|---|---|---|---|---|
| 33 | 4 | 3 | 0.75 | Jail |
| 34 | 4 | 4 | 1.00 | Jail |
| 35 | 5 | 5 | 1.00 | Jail |
| 36 | 6 | 1 | 0.17 | Jail |
| 37 | 6 | 6 | 1.00 | Jail |
| 38 | 9 | 9 | 1.00 | Jail |
| 39 | 10 | 8 | 0.80 | Jail |
| 40 | 13 | 12 | 0.92 | Jail |
| 41 | 2 | 0 | 0.00 | Community |
| 42 | 2 | 0 | 0.00 | Community |
| 43 | 2 | 0 | 0.00 | Community |
| 44 | 2 | 0 | 0.00 | Community |
| 45 | 2 | 0 | 0.00 | Community |
| 46 | 2 | 0 | 0.00 | Community |
| 47 | 2 | 0 | 0.00 | Community |
| 48 | 2 | 0 | 0.00 | Community |
| 49 | 2 | 0 | 0.00 | Community |
| 50 | 2 | 0 | 0.00 | Community |
| 51 | 2 | 0 | 0.00 | Community |
| 52 | 2 | 0 | 0.00 | Community |
| 53 | 2 | 0 | 0.00 | Community |
| 54 | 2 | 0 | 0.00 | Community |
| 55 | 2 | 0 | 0.00 | Community |
| 56 | 2 | 0 | 0.00 | Community |
| 57 | 2 | 0 | 0.00 | Community |
| 58 | 2 | 0 | 0.00 | Community |
| 59 | 2 | 0 | 0.00 | Community |
| 60 | 2 | 0 | 0.00 | Community |
| 61 | 2 | 0 | 0.00 | Community |
| 62 | 2 | 0 | 0.00 | Community |
| 63 | 2 | 0 | 0.00 | Community |
| 64 | 2 | 0 | 0.00 | Community |
| 65 | 2 | 0 | 0.00 | Community |
| 66 | 2 | 0 | 0.00 | Community |
| 67 | 2 | 0 | 0.00 | Community |
| 68 | 2 | 0 | 0.00 | Community |
| 69 | 2 | 0 | 0.00 | Community |
| 70 | 2 | 0 | 0.00 | Community |
| 71 | 2 | 0 | 0.00 | Community |

| | | | | |
|---|---|---|---|---|
| 72 | 2 | 0 | 0.00 | Community |
| 73 | 2 | 0 | 0.00 | Community |
| 74 | 2 | 0 | 0.00 | Community |
| 75 | 2 | 0 | 0.00 | Community |
| 76 | 2 | 0 | 0.00 | Community |
| 77 | 2 | 0 | 0.00 | Community |
| 78 | 2 | 0 | 0.00 | Community |
| 79 | 2 | 0 | 0.00 | Community |
| 80 | 2 | 0 | 0.00 | Community |
| 81 | 2 | 0 | 0.00 | Community |
| 82 | 2 | 1 | 0.50 | Community |
| 83 | 2 | 1 | 0.50 | Community |
| 84 | 2 | 1 | 0.50 | Community |
| 85 | 2 | 1 | 0.50 | Community |
| 86 | 2 | 1 | 0.50 | Community |
| 87 | 2 | 2 | 1.00 | Community |
| 88 | 2 | 2 | 1.00 | Community |
| 89 | 2 | 2 | 1.00 | Community |
| 90 | 2 | 2 | 1.00 | Community |
| 91 | 2 | 2 | 1.00 | Community |
| 92 | 2 | 2 | 1.00 | Community |
| 93 | 2 | 2 | 1.00 | Community |
| 94 | 2 | 2 | 1.00 | Community |
| 95 | 2 | 2 | 1.00 | Community |
| 96 | 2 | 2 | 1.00 | Community |
| 97 | 2 | 2 | 1.00 | Community |
| 98 | 2 | 2 | 1.00 | Community |
| 99 | 3 | 0 | 0.00 | Community |
| 100 | 3 | 0 | 0.00 | Community |
| 101 | 3 | 0 | 0.00 | Community |
| 102 | 3 | 0 | 0.00 | Community |
| 103 | 3 | 0 | 0.00 | Community |
| 104 | 3 | 0 | 0.00 | Community |
| 105 | 3 | 0 | 0.00 | Community |
| 106 | 3 | 3 | 1.00 | Community |
| 107 | 3 | 3 | 1.00 | Community |
| 108 | 3 | 3 | 1.00 | Community |
| 109 | 3 | 3 | 1.00 | Community |
| 110 | 4 | 0 | 0.00 | Community |

| | | | | |
|---|---|---|---|---|
| 111 | 4 | 2 | 0.50 | Community |
| 112 | 4 | 4 | 1.00 | Community |
| 113 | 5 | 4 | 0.80 | Community |
| 114 | 5 | 5 | 1.00 | Community |
| 115 | 6 | 4 | 0.67 | Community |
| 116 | 6 | 6 | 1.00 | Community |
| 117 | 8 | 8 | 1.00 | Community |
| 118 | 23 | 3 | 0.13 | Community |

**Supplemental Table S4.** Epidemiologic factors associated with *ermC* plasmid in jail-onset infections

| Characteristic | ***ermC* Plasmid Absent**, N = 66[1] | ***ermC* Plasmid Present**, N = 80[1] | **p-value**[2] |
|---|---|---|---|
| Race/Ethnicity | | | 0.8 |
| Hispanic/Latino | 6 (9.1%) | 10 (13%) | |
| Non-Hispanic Black | 41 (62%) | 50 (63%) | |
| Non-Hispanic White | 17 (26%) | 19 (24%) | |
| Other/Unable to Determine | 2 (3.0%) | 1 (1.3%) | |
| Sex | | | 0.033 |
| Female | 8 (12%) | 21 (26%) | |
| Male | 58 (88%) | 59 (74%) | |
| Year | | | 0.2 |
| 2015 | 11 (17%) | 9 (11%) | |
| 2016 | 28 (42%) | 45 (56%) | |
| 2017 | 27 (41%) | 26 (33%) | |
| Wound Infection | 64 (97%) | 78 (98%) | 0.8 |
| Recent cocaine use (past year)[3] | 5 (7.6%) | 7 (8.8%) | 0.9 |
| Recent heroin use (past year)[3] | 12 (18%) | 12 (15%) | 0.8 |
| Recent marijuana use (past year)[3] | 14 (21%) | 24 (30%) | 0.5 |
| Injection drug use (ever)[3] | 6 (9.1%) | 7 (8.8%) | >0.9 |
| Previous incarceration (past year) | 45 (68%) | 43 (54%) | 0.076 |
| Any clindamycin use[4, 5] | 6 (9.1%) | 25 (31%) | 0.001 |
| Macrolides use[5] | 6(9.1%) | 13 (16%) | 0.2 |

[1]n (%)

[2]Fisher's exact test; Pearson's Chi-squared test (all two-sided)

[3]Missing data for two cases (one *ermC*-present and one *ermC*-absent)

[4] Includes both clindamycin and clindamycin topical. Topical clindamycin was independently associated with *ermC* carriage (12% exposure with *ermC* versus 1.5% without *ermC,* p = 0.013), with only two patients receiving both topical and other forms of clindamycin.

[5] Antibiotic use was restricted to exposures at least 10 days and no more than 6 months prior clinical isolate collection

**Supplemental Table S5.** Epidemiologic factors associated with *ermC* plasmid in detainees colonized at intake

| Characteristic | *ermC* Plasmid Absent, N = 127[1] | *ermC* Plasmid Present, N = 20[1] | p-value[2] |
|---|---|---|---|
| Age | 34 (26, 49) | 37 (31, 46) | 0.6 |
| Race/Ethnicity | | | >0.9 |
| Hispanic/Latino | 15 (12%) | 3 (15%) | |
| Multiple | 1 (0.8%) | 0 (0%) | |
| Non-Hispanic Black | 95 (75%) | 15 (75%) | |
| Non-Hispanic White | 16 (13%) | 2 (10%) | |
| Sex | | | 0.2 |
| Female | 43 (34%) | 4 (20%) | |
| Male | 84 (66%) | 16 (80%) | |
| Year | | | 0.090 |
| 2016 | 29 (23%) | 9 (45%) | |
| 2017 | 84 (66%) | 9 (45%) | |
| 2018 | 14 (11%) | 2 (10%) | |
| Colonization site | | | 0.6 |
| Groin | 25 (20%) | 2 (10%) | |
| Nose | 68 (54%) | 13 (65%) | |
| Throat | 34 (27%) | 5 (25%) | |
| HIV | 53 (42%) | 11 (55%) | 0.3 |
| Recent cocaine use (past year)[3] | 52 (41%) | 12 (60%) | 0.12 |
| Recent heroin use (past year)[3] | 31 (25%) | 11 (55%) | 0.005 |
| Recent marijuana use (past year)[3] | 84 (67%) | 8 (40%) | 0.022 |
| Used needle for drugs (past year) | 17 (13%) | 6 (30%) | 0.090 |
| Been to jail before | 113 (89%) | 19 (95%) | 0.7 |
| Been to jail before (more than 5 times) | 62 (49%) | 11 (58%) | 0.5 |
| Released from jail in past 6 months | 35 (28%) | 9 (45%) | 0.12 |
| Lived in substance abuse center (past year) | 18 (14%) | 6 (30%) | 0.10 |
| Lived in homeless shelter (past year) | 16 (13%) | 5 (25%) | 0.2 |
| Was homeless (past year) | 34 (27%) | 10 (50%) | 0.035 |
| Been to emergency room (past year) | 74 (58%) | 12 (60%) | 0.9 |
| Hospitalized (past year) | 36 (28%) | 5 (25%) | 0.8 |

[1]Median (IQR); n (%)

[2]Wilcoxon rank sum test; Fisher's exact test; Pearson's Chi-squared test (all two-sided)
[3]Missing data for one *ermC*-absent case

**Supplemental Table S6.** Demographic and clinical characteristics of MRSA cases in the community. This study included data from two sample periods of Cook County Health patients: Study A (2011-2014) and Study B (2004-2010; 2015-2020).

| Characteristic | **Study A**, N = 774[1] | **Study B**, N = 1,697[1] |
|---|---|---|
| Race/ethnicity | | |
| Non-Hispanic Asian | 12 (1.6%) | 33 (1.9%) |
| Non-Hispanic Black | 451 (59%) | 993 (59%) |
| Hispanic/Latino | 169 (22%) | 357 (21%) |
| Non-Hispanic White | 120 (16%) | 260 (15%) |
| Other | 14 (1.8%) | 53 (3.1%) |
| Unknown | 8 | 1 |
| Sex | | |
| Female | 221 (29%) | 579 (34%) |
| Male | 553 (71%) | 1,116 (66%) |
| Age | 43 (29, 52) | 44 (30, 55) |
| Year | | |
| 2004 | 0 (0%) | 57 (3.4%) |
| 2005 | 0 (0%) | 60 (3.5%) |
| 2006 | 0 (0%) | 62 (3.7%) |
| 2007 | 0 (0%) | 64 (3.8%) |
| 2008 | 0 (0%) | 65 (3.8%) |
| 2009 | 0 (0%) | 797 (47%) |
| 2010 | 0 (0%) | 99 (5.8%) |
| 2011 | 244 (32%) | 0 (0%) |
| 2012 | 189 (24%) | 0 (0%) |
| 2013 | 234 (30%) | 0 (0%) |
| 2014 | 107 (14%) | 0 (0%) |
| 2015 | 0 (0%) | 47 (2.8%) |
| 2016 | 0 (0%) | 35 (2.1%) |
| 2017 | 0 (0%) | 25 (1.5%) |
| 2018 | 0 (0%) | 22 (1.3%) |
| 2019 | 0 (0%) | 221 (13%) |
| 2020 | 0 (0%) | 143 (8.4%) |
| Infection Type | | |
| Blood | 41 (5.3%) | 184 (11%) |
| Respiratory | 36 (4.7%) | 75 (4.4%) |
| Urine | 6 (0.8%) | 60 (3.5%) |
| Wound | 657 (85%) | 1,320 (78%) |
| Other | 29 (3.8%) | 58 (3.4%) |

| **Characteristic** | **Study A**, N = 774[1] | **Study B**, N = 1,697[1] |
|---|---|---|
| Unknown | 5 | 0 |
| Onset | | |
| Community associated | 461 (60%) | 892 (53%) |
| Community onset, hospital associated | 220 (29%) | 611 (36%) |
| Hospital onset | 88 (11%) | 194 (11%) |
| Unknown | 5 | 0 |
| HIV-positive | 66 (8.5%) | 153 (9.0%) |
| Hospitalization in the last year | 159 (21%) | 445 (26%) |
| Emergency department visit in the last year | 325 (42%) | 945 (56%) |
| Ever incarcerated | 153 (20%) | 299 (18%) |
| Incarcerated in the last year | 72 (9.4%) | 125 (7.4%) |
| Unknown | 5 | 0 |
| Homeless, living on the streets currently | 65 (8.5%) | 71 (5.2%) |
| Unknown | 11 | 333 |
| Drug use ever | 392 (53%) | 496 (36%) |
| Unknown | 41 | 327 |
| ErmC+ | 144 (19%) | 219 (13%) |
| Unknown | 0 | 58 |

[1]n (%); Median (IQR)

**Supplemental Table S7. Accessions and meta-data for genomes included in this study.**

*See attached spreadsheet.*
